# Entrepreneurial Methods for Healthcare Redesign

**DOI:** 10.64898/2026.07.28.26358976

**Authors:** Jacob Howran, Aryaman Sharma, Kaytlin Andrews, Dana Pople-McCord, Sabra K. Salim, David Jenka, Fabio Ynoe de Moraes, Katie Goldie, Corinne Babiolakis, Ryan Alkins, Shervin Taslimi, Chris Pasarikovski, Julius Ebinu, Douglas J. Cook, Ron Levy, James Purzner, Teresa Purzner

## Abstract

**Objective:** To evaluate whether entrepreneurial methodologies applied to system-wide healthcare redesign were associated with improved survival, care timeliness, and rural-urban equity among patients with glioblastoma.

**Design:** Non-randomized pre-post cohort study

**Setting:** Tertiary neuro-oncology centre in Ontario, Canada

**Participants:** Adults aged ≥18 years with histologically confirmed glioblastoma who underwent surgical resection between January 1, 2018, and February 28, 2025

**Interventions:** Implementation of the Integrative Brain Tumor Program (IBTP), a system-wide care intervention grounded in user-defined priorities and developed using a design thinking approach (empathize, define, ideate, prototype, test) integrated with operational frameworks adapted from early-stage innovation. System change was treated as a deliberate, deployable intervention that could be designed, launched, iteratively refined, and evaluated. Coordinated changes were embedded across healthcare services within existing infrastructure and resource constraints through a single centralized nurse navigator who standardized referrals, patient education, and real-time care coordination.

**Main Outcomes and Measures:** Primary outcomes were one-year overall survival and time to postoperative MRI completion and radiotherapy initiation. Secondary outcomes assessed rural-urban equity in these measures. Associations were evaluated using Cox proportional hazards and Fine-Gray competing-risk models adjusted for age, sex, rurality, MGMT promoter methylation status, and calendar time.

**Results:** Among 297 patients (244 pre-implementation, 53 post-implementation), baseline demographic and tumor characteristics were similar across cohorts. One-year overall survival was higher in the post-implementation cohort (60.4% vs 42.2%), corresponding to an adjusted hazard ratio of 0.61 (95% CI, 0.38–0.99). Postoperative MRI completion within 48 hours increased from 45.9% to 66.0% (adjusted cause-specific hazard ratio, 1.50; 95% CI, 1.05–2.14) with similar improvements observed at 7 days. Time to radiotherapy initiation did not differ between cohorts. Survival and MRI timeliness did not differ by rural or urban residence in either period, though rural radiotherapy delays were attenuated postimplementation.

**Conclusions:** Systematic application of entrepreneurial methods to health system redesign was associated with clinically meaningful improvements in glioblastoma survival and care timeliness using minimal resources (single nurse navigator). These findings suggest that treating system change as an intervention grounded in user-defined priorities and oriented toward integrated systems rather than sequential process optimization can support sustainable transformation of complex, coordination-dependent care pathways and warrants evaluation in other disease settings.

**What is already known on this topic:** Glioblastoma outcomes remain poor, and prior improvement efforts have largely targeted isolated process measures rather than integrated system-level redesign. Design thinking has been described in healthcare innovation, but few studies have evaluated its impact on survival or equity in oncology, and rural–urban disparities in glioblastoma care persist.

**What this study adds:** In a quasi-experimental cohort study, system-wide redesign using design thinking and entrepreneurial methods was associated with higher one-year survival and faster postoperative MRI completion without changes to oncologic therapy or added infrastructure. Rural disadvantages in radiotherapy initiation were attenuated after implementation.

**How this study might affect research, practice, or policy:** Treating system change as a structured, stakeholder-driven intervention may improve timeliness, survival, and equity in complex cancer pathways. Similar low-resource, navigator-led models warrant evaluation in other disease settings.

## INTRODUCTION

Healthcare systems, across both public and private models, are increasingly strained by widening disparities in care access, quality, and outcomes.^1–3^ As health systems serve increasingly diverse populations across urban, rural, socioeconomic, and cultural contexts, there is a growing need for approaches that are adaptive, integrated, and responsive to patient and provider experience^1,3,4^. One unconventional but powerful source of such approaches lies beyond academic and clinical traditions: the entrepreneurial sector. Early-stage companies operate in high-stakes environments where continued viability depends on rapidly identifying unmet needs, designing broadly applicable solutions, and iteratively refining them to meet user demands. Among the methodologies developed in this context, design thinking offers a structured approach to innovation that prioritizes user-defined needs, iterative prototyping, and continuous stakeholder feedback^5,6^. Although originally developed for product design rather than healthcare delivery, design thinking provides a framework that can be intentionally applied to health systems to elicit system-level priorities from patients and practitioners, co-develop coordinated interventions that integrate within existing organizational and resource constraints, and prospectively track intervention performance to enable real-time refinement based on user feedback.

Design thinking differs from traditional quality improvement in two critical ways: first, it begins with patient- and provider-identified barriers rather than institutionally or investigator-defined assumptions; and second, it embeds rapid prototyping with continuous stakeholder input. Critically, this approach is oriented toward redesigning the care pathway as an integrated system rather than sequentially optimizing individual processes in isolation^7^. In this way, design thinking enables coordinated change across interdependent services and complements traditional quality improvement by aligning interventions with user-defined needs while supporting adaptation in complex clinical environments.

Glioblastoma (GB), the most common and aggressive primary malignant brain tumour in adults, offers an especially sensitive model for testing design thinking as an approach to system-level innovation^8,9^. GB is classified as a World Health Organization (WHO) 2021 Grade IV glioma and affects approximately 3 per 100,000 people in North America, with a median age at diagnosis of 64 years.^10^ Despite decades of research, outcomes remain poor: treatment-free survival is typically 3-4 months, median overall survival is approximately 15 months, and five-year survival is approximately 5%.^9^ Because disease progression is rapid and outcomes are closely tied to timely intervention, even small inefficiencies or delays in care delivery can have a significant impact on survival and quality of life. In this context, addressing coordination failures one process at a time risks prolonged implementation timelines and potentially diminishing efficacy of earlier interventions, underscoring the need for system-wide redesign rather than sequential process optimization. Prior efforts to improve glioblastoma care have focused largely on isolated process measures, such as imaging timeliness or referral efficiency, with limited evaluation of integrated system-level redesign or downstream clinical outcomes.^3,11,12^ Although design thinking and other entrepreneurial methodologies have been explored in health services innovation, their application to complex care pathway redesign, particularly in oncology, has been largely descriptive and rarely linked to survival or equity endpoints. Moreover, few studies have examined whether care-pathway redesign can mitigate rural–urban disparities in GB outcomes within a centralized referral system. We therefore applied a design thinking framework grounded in user-defined priorities to redesign a regional, system-wide neuro-oncology care pathway, treating system change itself as the unit of intervention, and prospectively evaluated temporal associations with survival, care timeliness, and geographic equity.

## METHODS

### Study Overview

To assess whether the Integrative Brain Tumour Program (IBTP) was associated with differences in outcomes and equity metrics in GB care, we conducted a non-randomized, pre-post cohort study using linked clinical and operational data. This quasi-experimental design reflects the system-level nature of the intervention, for which individual randomization was not feasible. We applied advanced survival and competing-risk methods to adjust for baseline imbalances and secular trends inherent to this quasi-experimental design (Supplementary Methods S1 and S4).

The IBTP comprised of five integrated components designed to restructure early postoperative neuro-oncology care: (1) a centralized nurse navigator, (2) standardized milestone-driven referral pathways, (3) real-time referral tracking and escalation, (4) structured patient and caregiver education, and (5) coordinated regional provider engagement. These components were implemented concurrently as a system-wide intervention rather than as isolated process changes and were developed through iterative stakeholder co-design.

The intervention was designed to address three failure points commonly observed in GB care: delayed coordination across disciplines, lack of real-time accountability for missed milestones, and limited patient-facing transparency regarding care progression. By centralizing navigation, standardizing referral milestones, and enabling active tracking and escalation, the program aimed to reduce time-dependent attrition and fragmentation during the early postoperative period, when downstream treatment decisions are most sensitive to delay. Any association with survival was hypothesized to occur indirectly through earlier treatment initiation, reduced care discontinuity, and improved adherence to evidence-based pathways rather than through changes in therapeutic modality.

### Intervention Development

#### Framework and Process Overview

The IBTP was developed using iterative design thinking modes adapted from the Stanford d.school (Hasso Plattner Institute of Design at Stanford). These modes (empathize, define, ideate, prototype, and test) were applied in a flexible, non-linear manner to systematically identify system-level barriers, prioritize intervention targets, and refine solutions through repeated stakeholder feedback. Detailed qualitative methods, stakeholder composition, co-design activities, and iterative prototyping outputs are described in the Supplementary Appendix (Supplementary Methods S2; Supplementary Tables 1–4; Supplementary Figures 2–9). Figure 1 summarizes the objectives, inputs, and outputs from each design stage.

**Figure 1.**
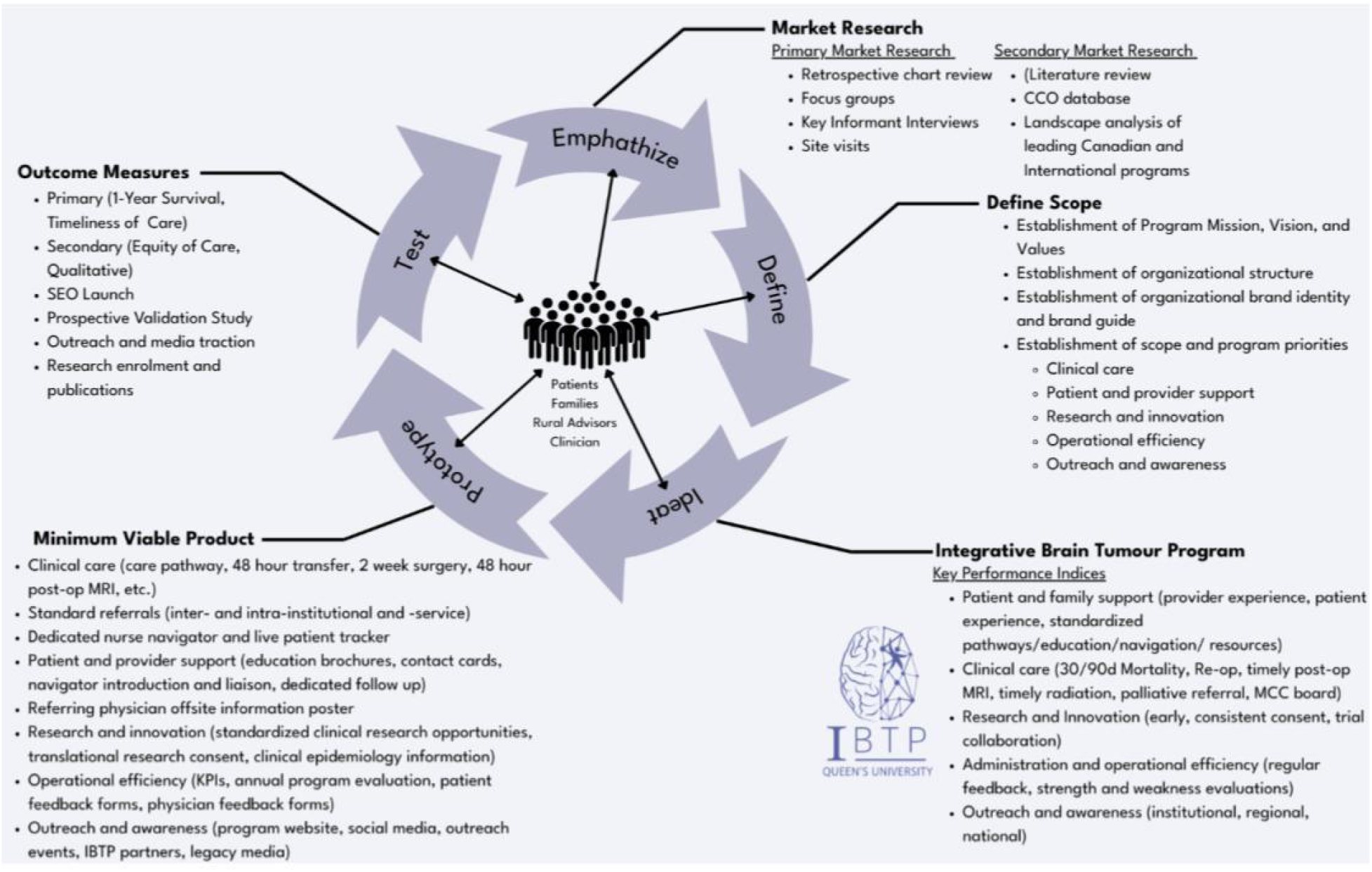
Design Thinking Framework and Stakeholder Engagement. The model illustrates the human-centered approach of the IBTP, emphasizing the participation of patients, families, and providers throughout all stages of co-development. Grounded in the Stanford d.School design thinking models, the cycle proceeded through empathize, define, ideate, prototype, and test stages. Primary and secondary market research, including retrospective chart reviews, interviews, and institutional benchmarking, identified systemic gaps in communication, care coordination, and equity. These insights informed scope definition, emphasizing standardized referrals, early palliative and psychosocial support, and rural and Indigenous inclusion. The resulting minimum viable product (MVP) included a nurse navigator, live referral tracking tools, and educational resources. Program outcomes were evaluated using key performance indices (KPIs) encompassing survival, timeliness, equity, patient experience, and system efficiency. This structured approach enabled the rapid translation of patient and provider insights into a scalable care model. Additional design thinking processes are described in Supplementary Tables 1-4 which describe insights from a PESTLE analysis, SWOT analysis, and Value Proposition Canvas.

**Figure 2.**
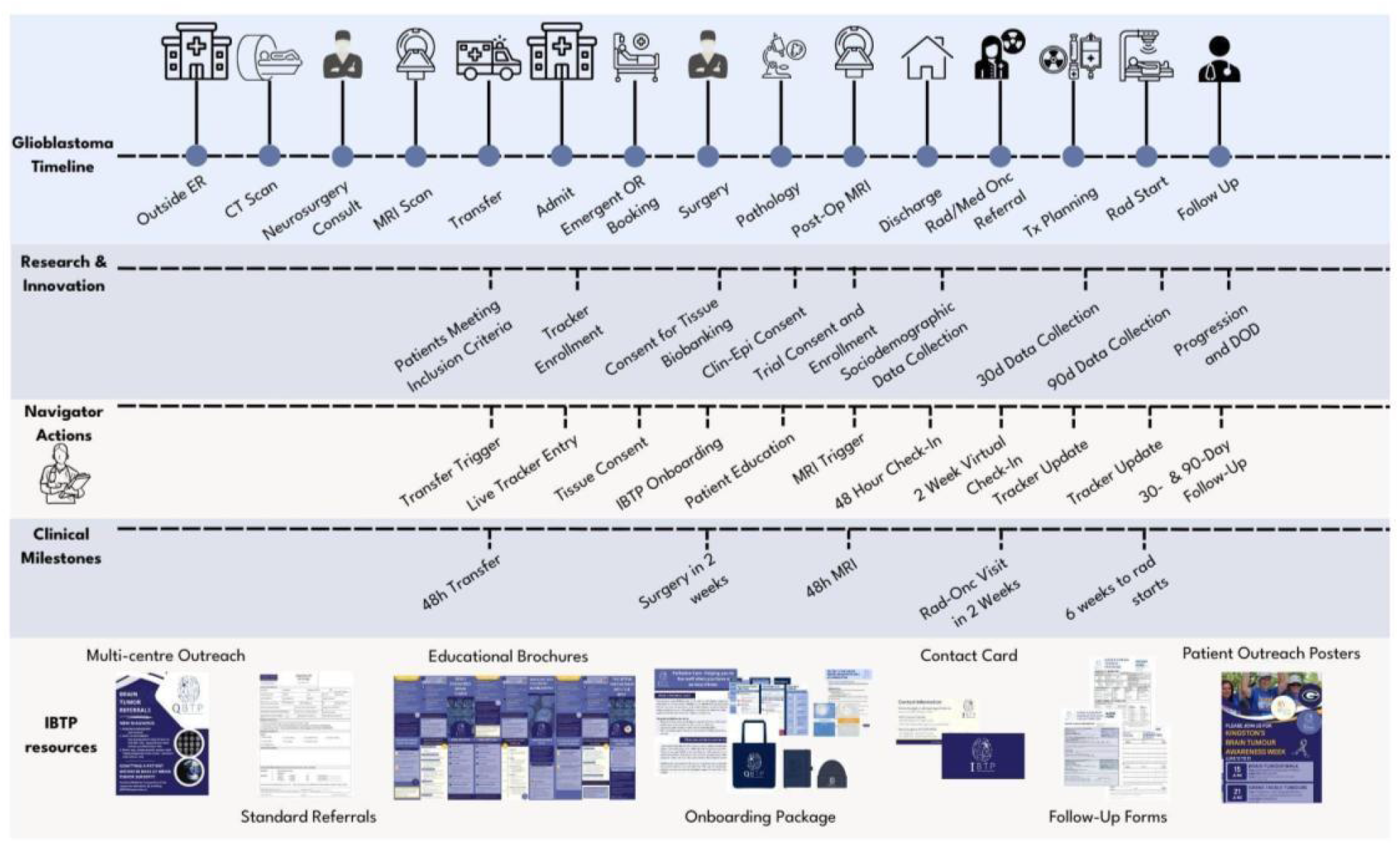
Integrative Brain Tumor Program (IBTP) Timeline of Care and Embedded Interventions. Clinical journey map developed through design-thinking methods, illustrating how patient and provider experiences were translated into an integrated care pathway. Journey mapping, a core design-thinking tool, enabled identification of transition gaps and workflow pain points. The upper track outlines standard clinical milestones, including neuroimaging, surgical intervention, oncology referral, and treatment initiation. The lower track details targeted IBTP interventions aligned to each phase, including standardized referral forms, early navigator contact, accelerated imaging protocols, automated referrals, and structured follow-up. Embedded throughout are research-enabling processes, such as consent, biospecimen collection, and longitudinal clinical data capture, facilitating translational science without disrupting clinical flow. The IBTP model operationalizes system-wide redesign through coordinated navigator-led support, institutional outreach, and real-time integration of clinical and research pathways. Sample cases describing a patient and provider experience navigating the glioblastoma care pathway pre- and post-IBTP are detailed in Supplementary Tables 5 and 6.

**Figure 3.**
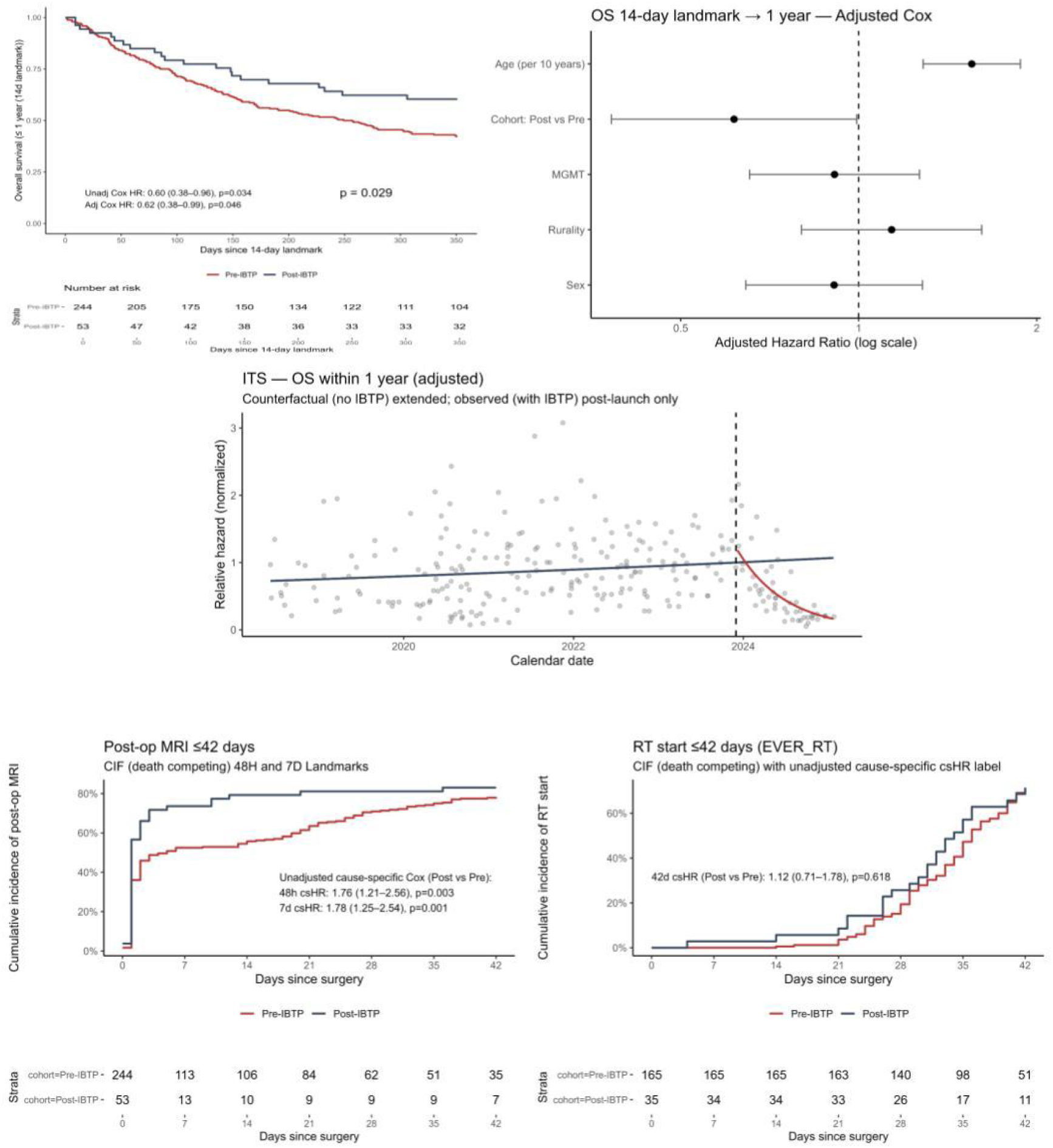
One-year Overall Survival and Timeliness Cumulative Incidence Functions for Postoperative MRI and Radiotherapy Initiation Pre- and Post-Integrative Brain Tumour Program (IBTP) (Top Left) Kaplan-Meier survival curves show higher 1-year survival in the post-IBTP period (HR 0.60, 95% CI 0.38-0.96; p = 0.034), and adjusted cox models estimate lower hazard of death after implementation. Numbers at risk are shown below the x-axis. The adjusted Cox proportional-hazards model (Top Right) includes age, sex, rurality, and MGMT promoter methylation status. Post-IBTP status remained independently associated with lower mortality risk after multivariable adjustment. Error bars represent 95% confidence intervals on a log scale. Interrupted time-series analyses (Middle) showed lower post-implementation hazard estimates compared with model-based counterfactual projections from pre-intervention trends. Aalen’s cumulative effect of calendar time reveals a modest increase in mortality to 0.06 by IBTP launch indicating a modest increase in mortality prior to launch, further attenuating the impact of secular trend bias. The post-IBTP period was associated with faster completion of postoperative MRI (Bottom Left; cause-specific HR 1.55, 95% CI 1.04-2.31; p = 0.041; subdistribution HR 1.40, 1.01-1.95), whereas time to radiotherapy initiation was not significantly different between cohorts (Bottom Right; HR 1.04, 0.64-1.68; p = 0.88). Fine-Gray and calendar-time adjusted models demonstrated consistent directionality and stability of estimates, confirming a sustained, calendar-time-adjusted improvement in postoperative MRI timeliness following IBTP implementation, while radiotherapy start intervals remained unchanged.

**Figure 4.**
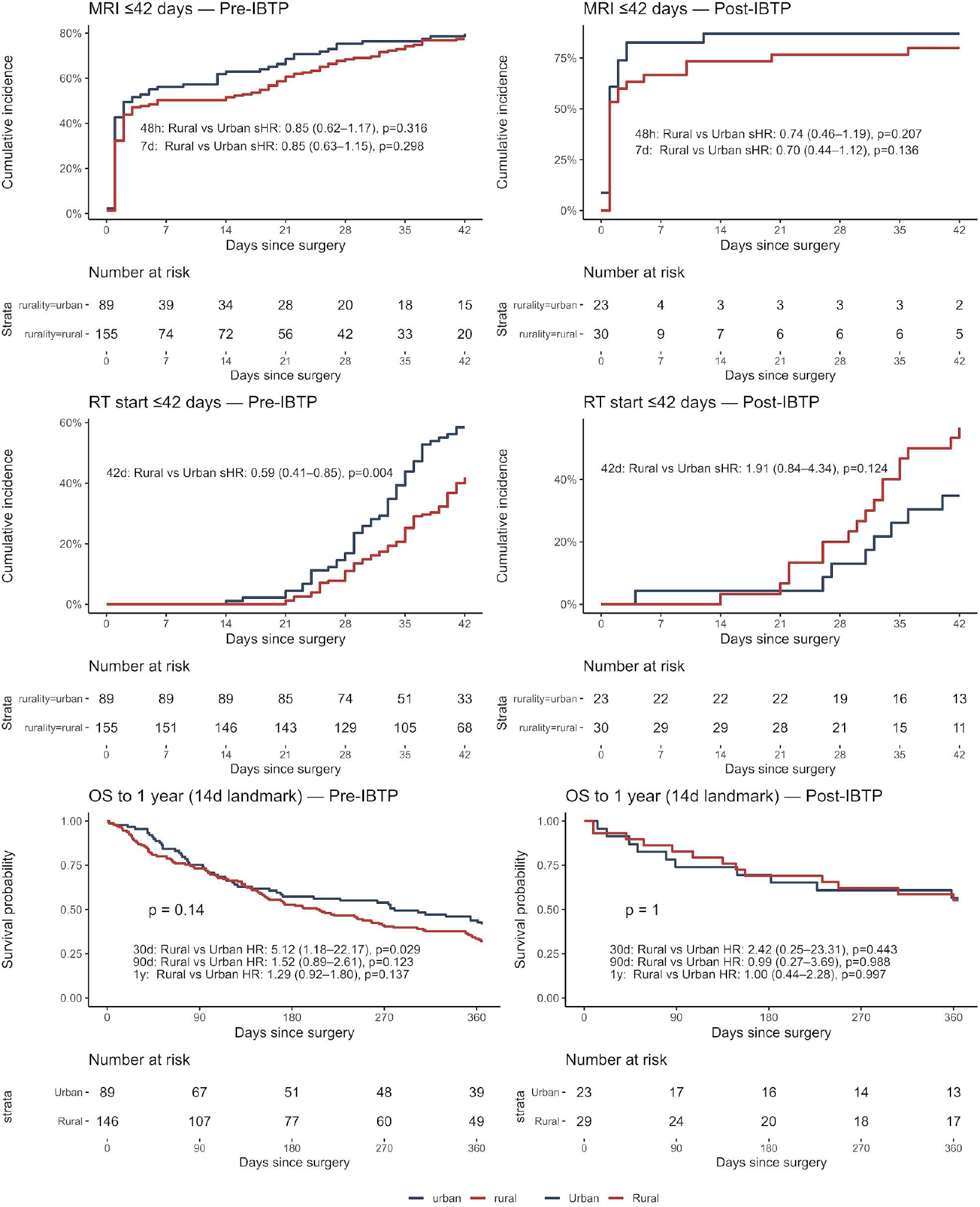
Urban-Rural Outcomes after Integrative Brain Tumor Program (IBTP) implementation. Urban–rural outcomes before and after Integrative Brain Tumour Program (IBTP) implementation are shown. Cumulative-incidence curves depict postoperative MRI completion within ≤42 days (top row) and radiotherapy (RT) start within ≤42 days among patients who received RT (middle row), stratified by rurality in the pre-IBTP (left) and post-IBTP (right) eras. MRI completion rates were similar between urban and rural patients in both eras, with no evidence of a rural–urban disparity. In contrast, rural patients experienced delayed RT initiation in the pre-IBTP era, a difference that was attenuated following IBTP implementation. Kaplan–Meier curves show one-year overall survival from a 14-day postoperative landmark (bottom row), with visibility lower survival among rural patients before IBTP and overlapping survival distributions after implementation. Numbers at risk are shown below each plot, and hazard ratios or subdistribution hazard ratios with 95% confidence intervals are annotated within panels. Subgroup sizes are small, and analyses were not powered for formal rurality-by-era interaction testing.

Using a structured design thinking framework, we conducted mixed-methods stakeholder engagement to identify system-level barriers and inform development of a regional neuro-oncology intervention. During the empathize and define stages, a retrospective chart review and 22 qualitative interviews with patients, caregivers, clinicians, and system leaders identified fragmented care transitions, delayed supportive services, and inconsistent communication (Supplementary Methods S2; Supplementary Table 1). Findings were benchmarked against national and international programs and validated through key-informant consultations. These insights informed a multidisciplinary co-design workshop that formalized program scope, mission, governance, and key performance indicators, with explicit prioritization based on feasibility within existing resources and anticipated impact on early postoperative coordination, followed by rapid prototyping and iterative deployment using agile feedback mechanisms.

The resulting IBTP consisted of standardized clinical pathways and referral tools, a centralized navigator-led coordination model with embedded milestone-based touchpoints, harmonized patient-facing educational resources and communication standards, and a regional outreach and awareness strategy (Supplementary Figures 2–9). Collectively, these components established a unified care architecture, reduced variability in referral timing, and created the operational and data infrastructure required for prospective monitoring of care timeliness and downstream evaluation of survival outcomes.

### Evaluation of Implementation Efficacy

#### Study design and participants

A non-randomised, mixed retrospective-prospective cohort study was performed at a single tertiary neuro-oncology centre in Ontario, Canada. Eligible participants were adults (aged ≥18 years) with WHO 2021-classified glioblastoma who underwent surgical resection at the study centre. The retrospective cohort included patients treated between Jan 1, 2018, and Nov 30, 2023, and the prospective cohort included patients treated between Dec 1, 2023, and Feb 28, 2025. Patients with prior glioma diagnoses, no surgical resection, or insufficient follow-up were excluded (Supplementary Figure 1).

#### Data collection

Clinical, pathological, and demographic data were extracted from institutional chart systems. Tumour classification followed WHO 2021 criteria, and survival was calculated from the date of first surgical resection to death. Rurality was defined using Statistics Canada census subdivisions (<50,000 population) (Supplementary Methods S3). Full variable definitions and data linkage procedures are detailed in the Supplementary Appendix (Supplementary Methods S1, S3, and S4).

#### Outcomes

Primary outcomes were prespecified time to postoperative MRI, time to radiotherapy initiation, and one-year overall survival. Secondary outcomes included assessment of rural-urban equity across these measures and descriptive evaluation of implementation, informed by prospective navigator and stakeholder feedback (Supplementary Methods S4; Supplementary Table 7).

#### Statistical analysis

Outcomes before and after IBTP implementation were compared using Cox proportional hazards models for survival outcomes and Fine–Gray competing-risk models for timeliness outcomes, with adjustment for age, sex, rurality, and O6-methylguanine-DNA methyltransferase (MGMT) promoter methylation status. Cause-specific hazards were estimated for timeliness outcomes to account for the competing risk of death. Proportional hazards assumptions were assessed using Schoenfeld residuals, and 14-day postoperative landmark analyses were prespecified to mitigate immortal time bias from early mortality. Calendar time was evaluated in sensitivity analyses using linear terms, restricted cubic splines with varying degrees of freedom, and interrupted time-series models. Flexible spline specifications were not selected as primary adjustments because of collinearity with intervention era and model instability. Prespecified sensitivity analyses included landmarking, competing-risk modeling, restriction to WHO 2021–classified tumors, death-date perturbation analyses, calendar-time adjustment, and bootstrap resampling. Temporal specificity of observed associations was further assessed using linear and nonlinear interrupted time-series models, including Aalen additive hazards models. Rural–urban equity evaluated using prespecified interaction terms (Supplementary Methods S4). All analyses were conducted using R statistical software.

#### Sex and Gender Reporting Statement

Analyses were disaggregated by recorded biological sex where feasible. Gender identity was not routinely collected in the clinical datasets used and could not be analysed separately. This limitation reflects current practices in routine neuro-oncology care and may obscure important gender-related differences in access, experience, and outcomes. Future work should prioritise systematic collection of sex and gender data to support more equity-informed analyses.

#### Patient and Public Involvement Statement

Patients and caregivers were not involved in the design or analysis of this specific evaluation study. However, patient and caregiver perspectives informed the development of the IBTP through prior needs assessments, qualitative feedback, and stakeholder engagement. These insights shaped the care processes and outcomes evaluated. Patients were not involved in data analysis or interpretation. Results will be shared with patient and caregiver communities through program dissemination activities.

## RESULTS

Outputs of the IBTP program, including implementation metrics and KPI-guided process measures, as well as associated robustness analyses, are reported in the Supplementary Appendix. The results below present prespecified clinical outcomes and equity analyses.

### Timeliness and Survival Outcomes

Outcomes were evaluated to test whether implementation of the IBTP care model, including centralized navigation and standardized referral milestones, was associated with differences in survival, care timeliness, and rural–urban equity. A total of 297 patients with GB were included, comprising 244 treated in the pre-IBTP era (January 1, 2018 to November 30, 2023) and 53 treated after implementation (December 1, 2023 to February 28, 2025). Baseline demographic, molecular, and geographic characteristics were well balanced between cohorts, supporting comparability at baseline (Table 1).

**Table 1.** Baseline demographic and tumour characteristics of patients treated before and after implementation of the Integrative Brain Tumour Program (IBTP) Data are presented as mean (SD) or n (%). Standardized mean difference (SMD) values quantify between-group balance, with 95% CIs shown in parentheses. Unknown MGMT and unclear extent-of-resection values were excluded from calculations. Unknown MGMT most often reflects patient tumours treated prior to standardization of methylation status assessment.

| Characteristic | Cohort |  | SMD<sup>*</sup> | 95% CI<sup>*</sup> |
| --- | --- | --- | --- | --- |
|  | Pre-IBTP n(%) | Post-IBTP n(%) |  |  |
| **Age (years)** |  |  | 0.00 | -0.29, 0.30 |
| Mean (SD) | 65.29 (11.65) | 65.26 (9.67) |  |  |
| **Sex** |  |  | 0.07 | -0.22, 0.37 |
| Female | 92 (38%) | 22 (42%) |  |  |
| Male | 151 (62%) | 31 (58%) |  |  |
| **MGMT status** |  |  | 0.56 | 0.26, 0.86 |
| Methylated | 113 (47%) | 25 (47%) |  |  |
| Unmethylated | 99 (41%) | 28 (53%) |  |  |
| Unknown | 31 (13%) | 0 (0%) |  |  |
| **Rurality** |  |  | 0.14 | -0.16, 0.44 |
| urban | 89 (37%) | 23 (43%) |  |  |
| rural | 154 (63%) | 30 (57%) |  |  |

<sup>\*</sup> Standardized Mean Difference; Abbreviation: CI = Confidence Interval

In unadjusted analyses, 1-year overall survival was higher in the post-IBTP cohort compared with the pre-IBTP cohort (60.4% vs 42.2%; hazard ratio [HR], 0.60; 95% CI, 0.38–0.96; P = .03). This association remained after adjustment for age, sex, rurality, MGMT promoter methylation status, and calendar year (adjusted HR, 0.61; 95% CI, 0.38–0.99; P = .046). Increasing age was independently associated with higher mortality per 10-year increase. Results were consistent in a 14-day postoperative landmark analysis (HR, 0.60; 95% CI, 0.38–0.96), indicating that the observed association was not driven by early perioperative mortality. Across prespecified sensitivity analyses, including WHO 2021 tumor reclassification, calendar-time spline adjustment, plus or minus 30-day perturbation of death dates, and bootstrap resampling, the direction and magnitude of the association were stable, with hazard ratios ranging from 0.55 to 0.60 and no evidence of violation of proportional hazards assumptions (Supplementary Methods S4; Supplementary Table 8). The absolute improvement in 1-year survival of approximately 18% occurred within a disease context characterized by rapid early progression and no contemporaneous changes in oncologic treatment protocols, supporting the plausibility of an association between care-pathway redesign and improved 1-year survival. Ninety-day survival was numerically higher in the post-IBTP cohort (83.0% vs 73.8%), although this difference did not reach statistical significance.

Post-IBTP patients experienced earlier postoperative MRI completion. The proportion receiving MRI within 48 hours increased from 45.9% to 66.0%, corresponding to faster MRI completion in adjusted cause-specific Cox models (hazard ratio, 1.50; 95% CI, 1.05–2.14). Similar improvements were observed for MRI completion within 7 days. Results were directionally consistent across competing-risk models, calendar-time–adjusted analyses, interrupted time-series models, and bootstrap resampling (Supplementary Methods S4; Supplementary Tables 8–9). Given the central role of early postoperative imaging in confirming extent of resection and guiding adjuvant treatment planning, these findings are consistent with improved perioperative coordination.

In contrast, time to radiotherapy initiation did not differ meaningfully between eras across adjusted, competing-risk, calendar-time–adjusted, or interrupted time-series analyses, with consistent null findings in sensitivity analyses (Supplementary Tables 8 and 10).

### Equity Outcomes

Because the IBTP did not include rural-specific interventions, rural-urban comparisons were evaluated as prespecified indicators of equity rather than direct targets of intervention. Before the IBTP implementation, rural patients had numerically lower 1-year survival, although differences were not statistically significant. After implementation, survival curves overlapped closely between rural and urban patients. Adjusted landmark and competing-risk models yielded consistent findings, supporting equity-neutral survival outcomes across eras (Supplementary Tables 8 and 10). Notably, early mortality (30-day) was significantly worse in rural patients compared to urban patients in the pre-IBTP cohort (HR 5.12, 95% CI 1.18-22.17, p=0.029) but this gap closed for the post-IBTP cohort. Visual inspection of the survival curves between urban and rural demonstrates nearly identical survival at 90-days with the largest gaps appearing in the early preoperative period (30 days) and later disease course (1 year).

Postoperative MRI timeliness did not differ by rural or urban residence before or after IBTP implementation. Adjusted cause-specific and competing-risk models confirmed the absence of geographic differences, indicating comparable access to postoperative MRI across regions in both eras. In contrast, radiotherapy initiation showed evidence of a pre-existing rural disadvantage. Before IBTP implementation, rural patients initiated radiotherapy later than urban patients. After implementation, this difference was no longer apparent, with overlapping initiation curves and directionally earlier radiotherapy among rural patients in adjusted models, although estimates were imprecise because of limited sample size. Collectively, these findings are consistent with attenuation of pre-existing rural-urban differences in radiotherapy initiation following IBTP implementation.

Overall, IBTP implementation was associated with higher one-year survival, faster postoperative MRI completion, and possible attenuation of pre-existing rural-urban differences in radiotherapy initiation.

## DISCUSSION

In this study, a neuro-oncology program was redesigned through a simple yet unconventional question: “what if we created a brain tumor program the way we would launch an early-stage startup?” Through entrepreneurial principles grounded in design thinking and KPI-guided iteration, the IBTP translated processes commonly used in early-stage innovation into neuro-oncology care. Implementation was associated with an approximately 40% lower hazard of death at one year, improvements in care timeliness, and attenuation of rural-urban differences, achieved within the first year, without changes to oncologic therapeutics or expansion of clinical infrastructure. Similar associations between care-delivery redesign and clinical outcomes have been observed in oncology, including randomized trials demonstrating that workflow-level interventions improving communication and monitoring can translate into clinically meaningful benefits, and pragmatic implementation trials evaluating system-level changes when individual randomization is infeasible ^11,13,14^.

In this study, design thinking did not replace conventional quality-improvement methods but provided a complementary structure for integrating data-driven analysis with iterative testing and real-world feedback. Retrospective analyses and qualitative interviews informed early priorities, while structured prototyping and revision allowed interventions to be refined in response to patient and provider experience. This adaptive approach enabled identification and correction of unintended consequences early in implementation, improving acceptability while maintaining operational feasibility.

A defining feature of this framework is its user-defined orientation. Rather than beginning with institutionally defined assumptions, design thinking foregrounds the experiences of patients, caregivers, and frontline providers across the care pathway. In this study, quantitative analyses identified delays and disparities, while qualitative stakeholder input clarified where coordination failures occurred and how they were experienced. Interventions were therefore designed to improve transparency, navigation, and communication rather than to introduce new clinical services, aligning system performance with how care is actually navigated. Notably, preservation of equity and numeric attenuation of rural-urban disparities occurred without targeted rural-specific interventions. When baseline inefficiencies were addressed through coordinated navigation and standardized workflows, equity gaps in radiotherapy narrowed as a consequence of inclusive system design rather than differential resource allocation. This finding supports the view that geographic disparities may reflect modifiable system-level failures rather than intrinsic differences in disease biology or access.

The generalizability of these findings may be supported by the program’s reliance on resource realignment rather than expansion; however, external validity is limited by the single-center setting and may vary across health systems with different referral structures, baseline timeliness, workforce models, and electronic infrastructure. The IBTP was implemented using a single $50,000 seed grant and did not involve additional clinical staffing. The nurse-navigator role was derived through repurposing and alignment of existing volume-based brain tumour funding, rather than creation of new funded positions. Importantly, this study does not compare design thinking with alternative quality-improvement approaches, nor does it establish superiority over conventional methods. Rather, it provides empirical validation that a design thinking, system-wide approach is feasible and can be associated with meaningful improvements in outcomes in a setting where no prior system-level optimization had been attempted. The observed associations are most likely to generalize to health systems characterized by centralized specialty care, shared electronic records, and identifiable coordination bottlenecks, where modest redesign can yield outsized gains. In fragmented or competitive delivery environments, analogous interventions may require additional governance or incentive alignment to achieve similar effects.

Several limitations warrant consideration. The study was non-randomized, and the post-implementation cohort was smaller, limiting power for subgroup and interaction analyses, particularly for equity endpoints. Residual confounding from unmeasured factors such as performance status or socioeconomic variables cannot be excluded. Findings derive from a single tertiary cancer centre with specific resources, operational structures and patient populations, necessitating caution to community practices and other jurisdictions. Follow-up was limited to one year, precluding assessment of longer-term outcomes, and although ITS and spline-adjusted models account for pre-intervention temporal trends, residual confounding due to other contemporaneous changes in care cannot be excluded. Retrospective data collection in the pre-implementation period may have introduced information bias, and COVID-19-related disruptions may have influenced baseline trends despite calendar-time adjustment suggesting otherwise. Despite these limitations, confidence in the observed associations is strengthened by consistency across multiple analytic approaches, including landmarking, competing-risk modelling, calendar-time adjustment, and sensitivity analyses. The temporal alignment between program implementation, process improvement, and outcome changes supports a plausible mechanistic link through improved coordination and timeliness of care (Supplementary Methods S1 and S4).

In summary, these findings suggest that stakeholder-driven, adaptive system redesign can meaningfully influence outcomes in complex oncology care. Future studies should assess reproducibility across other disease sites and health systems and evaluate longer-term survival and patient-reported outcomes.

## Conclusion

A stakeholder-driven, design thinking-informed approach to care-pathway redesign was associated with higher 1-year survival, faster postoperative imaging, and attenuation of geographic differences in glioblastoma care. These findings support the feasibility of entrepreneurial design frameworks as a viable strategy for system-level improvement in complex oncology settings and provide a proof of concept for treating health system change as an user-defined intervention that can be deliberately designed, launched, iteratively refined, and evaluated. Further study is needed to assess reproducibility across disease sites and health systems.

## Data Availability

All authors had full access to the data in the study and take responsibility for the integrity of the data and the accuracy of the data analysis. Deidentified participant data will be made available upon reasonable request to the corresponding author, subject to institutional and ethical approvals.

## Contributors

JH was the corresponding author and led data curation, formal analysis, software development, validation, visualization, and writing of the original draft, with supporting contributions to methodology. AS contributed equally to formal analysis and supported methodology, software, visualization, and drafting of the manuscript, and participated in manuscript review and editing. KA contributed to data curation, project administration, and drafting and revision of the manuscript. DPM contributed to conceptualization, led project administration, and contributed to data curation and supervision. SKS contributed to data curation, formal analysis, methodology, project administration, and manuscript review and editing. DJ contributed to conceptualization, methodology, validation, and manuscript review and editing. KG and CB contributed to conceptualization, methodology, project administration, and manuscript review and editing. FYM contributed to methodology and manuscript review and editing. RA, ST, CP, JE, RL, and DJC contributed to data curation, investigation, methodology, and manuscript review and editing. JP contributed to data curation, funding acquisition, resources, supervision, and manuscript review and editing. TP contributed equally to conceptualization, led methodology, project administration, and supervision, and contributed equally to manuscript review and editing.

The corresponding author attests that all listed authors meet authorship criteria and that no others meeting the criteria have been omitted. JH and TP act as guarantors of the work.

## Transparency Statement

The lead author (the manuscript’s guarantor) affirms that the manuscript is an honest, accurate, and transparent account of the study being reported; that no important aspects of the study have been omitted; and that any discrepancies from the study as originally planned have been explained.

## Role of the funding source

The funders of this study had no role in the study design, data collection, data analysis, data interpretation, or writing of the report. All authors had full access to the data and accepted responsibility for data integrity and accuracy.

## Declaration of interests

All authors declare no competing interests.

## Notes

### Competing Interest Statement

The authors have declared no competing interest.

### Author Declarations

The Health Sciences Research Ethics Board (HSREB) of Queen's University waived ethical approval for this work

