## Supplementary Methods for "Entrepreneurial Methods for Healthcare Redesign"

##### S1. Study Design and Analytic Framework

As the IBTP was undertaken as a complete system redesign wherein patients after launch are unable to be classified into non-IBTP intervention, a non-randomized pre-post quasi-experimental design was chosen. Secular trends were addressed using a calendar-time adjustment with cubic spline and trend changes were modeled using interrupted time series models. 14-Day landmark analysis was completed for all survival analyses to limit immortal time bias caused by early deaths. Fine-Gray competing risks analyses were completed for postoperative MRI and radiotherapy start including death as a competing risk, reported by subdistribution hazards (sHR) and cause-specific hazard ratios. For all outcomes, interrupted time-series (ITS) analysis was conducted to evaluate the slope trend in the pre-IBTP era and its immediate change at time-zero of IBTP launch.

##### S2. Intervention Development and Co-Design Process

Stakeholder engagement and qualitative research

During the empathize stage, we conducted both primary and secondary market research to identify patient and system-level barriers to care. Primary research included a retrospective chart review of glioblastoma patients treated in the region to map referral patterns, timelines, and care gaps prior to intervention. This quantitative analysis was paired with 22 qualitative interviews involving 43 participants, including patients, caregivers, rural advisors, physicians, and allied health professionals. Transcripts were analyzed thematically using an a priori coding framework independently applied by three researchers. Common themes included fragmented transitions of care, poor communication across settings, delayed access to palliative and psychosocial services, and lack of standardized educational materials. In addition, a targeted secondary market and landscape analysis was performed to benchmark against leading neuro-oncology programs in Canada and internationally. This analysis focused on structural design, patient pathway integration, navigation models, and research translation strategies. In parallel, key informant interviews were conducted with national and institutional system leaders, including the Director of the Canadian Cancer Clinical Trials Group and the heads of radiation and medical oncology at our institution. These consultations were used to identify operational bottlenecks, surface unmet needs, and align with institutional strategic priorities. Informed by principles common to early-stage startup development, this process enabled program development to be grounded in a clear understanding of existing market offerings, institutional readiness, and clinical feasibility.

Co-design and prioritization workshop

Importantly, each engagement activity in this study aligned with a corresponding design-thinking mode: interviews and journey mapping during the empathize stage, thematic synthesis during define, brainstorming and prioritization during ideate, and iterative pilot testing during prototype and test phases.

During the define and ideate stages, a multidisciplinary in-person stakeholder workshop was held with 34 participants to validate findings and co-prioritize intervention components. Workshop participants included neuropathology, neurosurgery, neuroradiology, neuro-oncology, patient advisors, community and cultural advisors, nursing, and allied health. Using real-time discussion and online polling, participants also formalized the program's mission, vision, and values, and defined the architectural structure of the regional program. Organizational roles were mapped, and personnel recruited to fulfill core positions. Program scope and priorities were established [Supplementary Table 1], key performance indicators (KPIs) were selected and benchmarked alongside provincially-defined targets to guide iterative evaluation and adaptation. Importantly,

Prototyping and iterative development

Program components, including patient navigation, standardized referral tools, community partnerships, and educational materials, were co-developed with the multidisciplinary patient and provider task force consulted during the prioritization workshop, and deployed during a structured "launch week" that included site visits to referring hospitals, institutional awareness campaigns, and community outreach. Early implementation was guided by an agile testing framework: prospective surveys, navigator field notes, and stakeholder feedback were used to identify early implementation barriers and power the next design cycle. This allowed real-time program refinement while supporting rapid scale and consistent service delivery across diverse rural and urban settings. This human-centered approach emphasized the lived experience of patients and providers (considering emotional, behavioral, and relational factors alongside operational efficiency) to ensure that redesigned workflows aligned with how people actually navigate care.

Program Development Outputs

Following the design and co-development phases, the intervention generated a comprehensive set of tools and structural components spanning clinical care, patient support, communication infrastructure, and regional engagement.

1. Program Architecture and Scope

The program generated a formalized mission, vision, and values statement and established the Integrated Brain Tumour Program (IBTP) across three divisions: (1) IBTP Clinical, which delivered patient care through standardized pathways and navigator-led coordination; (2) $IBT^2P$, which enabled embedded biospecimen collection, live-tracker registration, and prospective data capture for translational research; and (3) IBTP Outreach, which developed patient-facing educational resources and regional awareness campaigns (Figure 2). An organizational structure was created to support each domain, with multidisciplinary teams assembled from neurosurgery, neuro-oncology, neuroradiology, neuropathology, nursing, allied health, rural advisors, and Indigenous health representatives. Program architecture also codified KPIs into routine monitoring and evaluation [Supplemental Figure 1].

2. Standardized Pathways and Communication Tools

Standardized, milestone-driven care pathways were implemented to define (or align with provincially-defined) timelines for imaging, adjuvant therapy, and structured follow-up [Supplemental Figure 2].18 Referral processes were harmonized across the region through redesigned forms adopted both between referring institutions and KHSC, and internally within KHSC (e.g., neurosurgery to oncology) [Supplemental Figure 3]. These forms functioned as workflow triggers, automatically initiating processes such as registration for multidisciplinary case conferences (MCC). Scheduled referral checkpoints were embedded at diagnosis (to oncology), day 30 (to psychosocial care), and day 90 (to palliative care), ensuring consistent access to supportive services. Collectively, these outputs standardized patient flow across institutions, reduced variability in referral timing, and created the infrastructure that supported downstream evaluation of timeliness and survival outcomes (Figure 2).

3. Patient-Facing Resources

A comprehensive suite of educational and communication materials was produced and distributed to all patients following onboarding to the IBTP. These included brochures outlining care team roles, clinical language, milestone timelines, and contact information [Supplemental Figure 4], as well as handbooks from the Brain Tumour Foundation of Canada and logistical inserts (e.g., directions to hospital, parking details, palliative services) [Supplemental Figure 5]. Each patient received an introductory kit containing these resources along with branded items (tote bag, notebook, pen, hat) [Supplemental Figure 6]. Patients and caregivers were also provided with personalized contact cards for key team members to facilitate direct communication [Supplemental Figure 9]. To ensure consistency, all resources were standardized under a formal communication standardization campaign covering logos, fonts, colours, and tone. Collectively, these resources and outreach activities established a unified program identity, standardized communication, and enhanced patient engagement and awareness across the region.

4. Navigator Role and Patient Interface

A dedicated neuro-oncology patient navigator role was established and deployed as the central point of contact and care coordinator within the program. Using milestone-driven pathways, the navigator actively monitored progress, flagged at-risk cases, and ensured patients remained on schedule for treatment and follow-up. Standard touchpoints were embedded at initial diagnosis (often from an external center), preoperative orientation, postoperative introduction to the program, 48-hour post-discharge follow-up, and digital check-ins at two and four weeks. Formal navigator contact was repeated at 30 and 90 days to align with scheduled referral checkpoints. To support continuity of care, a brain-tumour-specific program email was created for non-urgent patient communication, and structured assessment and feedback forms were used to capture demographic, clinical, and experiential data [Supplemental Figure 7]. Navigator field notes and patient feedback were collected prospectively, providing real-time validation of implementation and identifying opportunities for iterative refinement. Collectively, the navigator role functioned as both a clinical coordination tool and a data capture interface, reinforcing patient support and operational efficiency within the program.

5. Regional Engagement and Outreach

The program was launched with an institutional awareness campaign and a structured "launch week," which included in-person visits to regional referring centers. Outreach materials were produced and disseminated to external providers, including brochures detailing referral pathways, contact cards, and wall posters outlining inpatient and outpatient referral criteria [Supplemental Figure 8]. Program visibility was reinforced through a coordinated communications strategy that included a program-specific website, an active social media presence (Instagram), and two features in regional newspapers. These activities established a recognizable program identity across the region, reinforced referral consistency, and ensured ongoing visibility among both providers and patients.

#### S3. Data Sources, Variable Definitions, and Cohort Construction

Surgical pathology was collected on all patients who underwent brain tumor surgery between April 1st, 2017, to March 31st, 2023, at KHSC. To reflect the 2021 CNS WHO classification, we have included only IDH WT GB, including high-grade variants such as gliosarcomas and giant-cell GB tumors with appropriate histological features (microvascular proliferation, necrosis, CDKN2A/B homozygous deletions, TERT mutant, EGFR amplification). Surgical pathology, date of birth (DOB), biological sex, fiscal year of diagnosis, date of operation, MGMT promoter methylation status per pathology report, extent or resection, and surgeon were collected on the patient chart system (PCS) affiliated with KHSC from oncology consult notes. Date of death (DOD) was determined using PCS death files and, when possible, cross-referenced with online obituaries. Age was calculated as years between DOB and operation. Survival was confirmed through clinical appointments and treatment consults. Patients who received multiple operations, survival was calculated based on the time between the first surgery and DOD.  Geographic information including home address, residing town, residing county, and postal code were obtained for all patients via PCS who met the inclusion criteria. The census subdivision associated with patient postal codes was determined using Statistics Canada 2021 census profile table. When postal codes corresponded with more than one census subdivision, the subdivision in closer geographic proximity to the patient address was used. Subdivisions with <50,000 residents were considered lower-populated areas, in keeping with provincially and federally recognized definitions of rurality in Canada and the United States. Importantly, this cutoff of 50,000 residents represents an inflection point in survival within the study catchment area, where sensitivity analyses in one of our previous studies identified patients in towns <50,000 as having a significantly worse survival.

Missing data in this study was largely found in the non-study variables. Age, sex, surgery date, radiation oncology visit/start for patients offered, and postoperative MRI for patients who received them, were all available. For patients who were not offered or who declined radiation, ever-RT vs all patient analysis was completed to remove the effect of patients who were never going to benefit from RT timeliness under the IBTP.  Patients whose MGMT methylation status were unknown because they were treated prior to the WHO 2021 reclassification model were still included in the analysis. Sensitivity analysis excluding all patients before 2021 was completed to remove possibly misclassified patients. Death date was ascertained through available hospital records and supplemented with obituaries when possible. Death-date-sensitivity analysis was conducted to minimize the ascertainment bias in this case. Patients whose postal codes did not match a consensus subdivision from the available division database were manually assessed, and when possible, the closest subdivision was used as a proxy for assignment. Inverse probability weighting was not appropriate in this case, as patient cohorts are temporally distinct and there is no possibility of cohort overlap, hence failing the positivity assumption in IPTW.

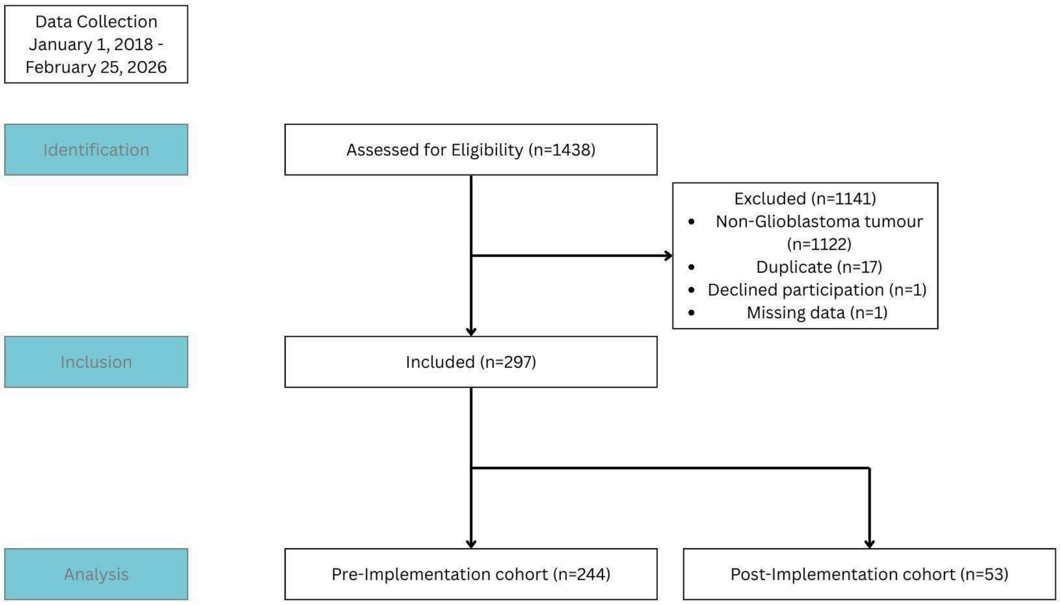

Supplemental Figure 1 **STROBE Flow Diagram of Cohort Assembly|** Flow diagram depicting identification, eligibility assessment, exclusions, and final cohort assembly for the study population. Between January 1, 2018, and February 25, 2026, a total of 1,438 patients were assessed for eligibility. Of these, 1,141 were excluded due to non-glioblastoma pathology, duplicate records, declined participation, or missing data. The final analytic cohort included 297 patients with histologically confirmed glioblastoma, stratified into a pre-implementation cohort (n=244) and a post-implementation cohort (n=53) based on the launch of the Integrative Brain Tumour Program.

#### S4. Statistical Models and Sensitivity Analyses

Survival at one year was assessed using Kaplan–Meier survival curves with between-group comparisons performed using log-rank tests. Unadjusted and adjusted Cox proportional hazards models were used to estimate hazard ratios (HRs) for mortality at 90 days and at one year. Multivariable models adjusted for age, sex, MGMT promoter methylation status, and rurality. To account for potential secular trends, calendar time was modeled using restricted cubic splines, allowing assessment of nonlinear secular trends independent of program implementation. Proportional hazards assumptions were evaluated using Schoenfeld residuals and complementary log(–log) survival plots. Timeliness outcomes were assessed using cumulative-incidence functions (CIFs), with between-group comparisons conducted using Gray’s test. Cause-specific Cox proportional hazards models were used to estimate HRs for postoperative MRI completion within

48 hours, 7 days, and 42 days, and for initiation of radiotherapy within 42 days. Competing-risk analyses were performed using Fine–Gray subdistribution hazard models, treating death as a competing event, and results are reported as subdistribution hazard ratios (sHRs). Rural equity was assessed using rurality-by-cohort interaction terms.

Bootstrap resampling was performed by sampling patients with replacement (n=2000). In each bootstrap replicate, the full model was refit and the statistic of interest recomputed. Confidence intervals were derived using the percentile bootstrap method. Bootstrap distributions were visualized using box and violin plots. Interrupted time-series (ITS) analyses were conducted to evaluate temporal changes in outcomes around the launch of the IBTP. Segmented Cox regression models were specified with an inflection at the IBTP implementation date, including terms for baseline temporal trend, an immediate post-implementation level change, and change in slope following implementation. Linear models were used for primary ITS analyses, with non-linear secular trends modeled in sensitivity analyses using natural cubic splines of calendar time. Model adequacy and temporal patterns were assessed through visual inspection of estimated hazard trajectories. Post-IBTP hazards of death were plotted against counterfactual projections derived from extrapolation of pre-intervention trends. To assess robustness, a bootstrap sensitivity analysis using random placebo intervention dates (n = 200) was performed to evaluate concordance with the true implementation date. Aalen's additive hazards models were used to further characterize pre-intervention trends and assess departures from proportionality over time.

To assess whether observed outcome differences could be explained by secular trends rather than by IBTP implementation, calendar time was evaluated using multiple Cox proportional hazards model specifications. Calendar time was modeled as a continuous variable using natural cubic splines with increasing degrees of freedom (df = 1–5), with internal knots placed at quantiles of the observed calendar-year distribution and boundary knots spanning the full study period. Model fit was evaluated using Akaike Information Criterion (AIC), stability of the cohort hazard ratio, and inspection of confidence intervals. Partial effect plots of the calendar-time spline term were generated to visualize secular trends. These demonstrated a weak, smooth trend without evidence of nonlinear inflection at the time of IBTP implementation; widening confidence intervals at the extremes reflected data sparsity rather than meaningful curvature. Restricted calendar-window analyses were additionally performed to reduce collinearity between cohort assignment and calendar time, and linear calendar-time terms and piecewise linear parameterizations were evaluated, yielding consistent results. Across all calendar-time specifications, increasing spline flexibility did not improve model fit and resulted in unstable cohort estimates due to collinearity between calendar time and intervention era. Because flexible calendar-time models absorbed intervention-related variation without improving explanatory power, they were treated as sensitivity analyses. Interrupted time-series Cox models, which explicitly parameterize baseline trends and post-implementation trajectory changes, were therefore selected as the primary analytic framework.

P-values <0.05 were deemed statistically significant. Given the hypothesis generating nature of secondary and sensitivity analyses, no formal adjustment for multiple comparison was applied. The authors acknowledge that conclusions may be drawn in cases of multiplicity from effect size and precision rather than strict p-value cutoffs.

### Supplementary Tables and Figures

#### Supplementary Tables

| Stage | Purpose | Actions |
| --- | --- | --- |
| **Empathize** | Gather a deep understanding of user needs through primary and secondary market research to develop a minimum viable product | • Primary Market Research<br>• Retrospective review of 1500 regional patients undergoing cranial surgery<br>• Focus groups with health care providers<br>• Qualitative interviews with patients, family members, healthcare administration and providers<br>• Secondary Market Research<br>• Landscape analysis of leading Canadian and international neuro-oncology programs<br>• Key informant interviews with national and institutional system leaders |
| **Define and Ideate** | Scope and priorities of the program, as well as the mission, vision and values are defined from insights gained during the empathize stage-informed by a multidisciplinary program leadership team consisting of brain tumour patient advisors, community and cultural advisors, physicians, nurses and allied health. Identified priorities are transformed into concrete, actionable outcomes | Establishment of Program Mission, Vision and Values<br>• Mission statement: To deliver the highest level of care and access to cutting-edge innovation for patients diagnosed with brain tumors in SEO through iterative, data-driven, and patient-centric care models.<br>• Vision Statement:To provide the highest level of care to every patient in Southeastern Ontario (SEO) diagnosed with a brain tumor, irrespective of where you are or who you are.<br>• Program Values: Collaboration, Equity, Data-Driven, Patient-Centred<br>Establishment of Organizational Structure<br>Establishment of IBTP Brand Identity and Brand Guide (Sfig X)<br>Establishment of Scope and Program Priorities:<br>• Clinical Care<br>o Improved 90 day and one year survival<br>o post-operative MRI within 48h<br>o initiation of radiation within 6 weeks<br>o Improved equity of care (rural vs urban patients)<br>o Early referrals and standardized access to palliative and psychosocial care<br>o Consistency of inclusion in multidisciplinary rounds<br>• Patient and Provider Support<br>o Early, consistent patient education<br>o Single patient touch point throughout journey<br>o Standardized, robust and efficient patient care pathway<br>• Research and Innovation<br>o early and consistent introduction of research opportunities to patients<br>o standardized tissue banking and established protocol for access to banked tissue<br>o collaboration with national neuro-oncology trials<br>• Operational Efficiency<br>o regular feedback on efficacy, strengths and weaknesses of the program from the perspective of patients and providers<br>• Outreach and Awareness<br>o establish program awareness at institutional, regional, provincial and national level |
| **Prototype** | | • Clinical Care<br>o Standardized care pathway with established milestones (Fig 1)<br>o transfers to tertiary care center within 48h of referral<br>o surgery within 2 weeks of diagnosis<br>o post-operative MRI within 48h<br>o first visit with radiation and medical oncology within 2 weeks of surgery<br>o Initiation of radiation within 6 weeks or surgery<br>• Standardized referrals |

| | | |
|---|---|---|
| | | <ul><li style="list-style:none"><ul><li>Standardized referrals from referring hospitals to tertiary care center, to ensure brain tumor patients are clearly differentiated from non-urgent referrals (Sfig 3)</li><li>Standardized referral from neurosurgery to radiation and medical oncology, which triggers automatic addition to multidisciplinary rounds (Sfig 3)</li></ul></li><li>Dedicated Nurse Navigator with Live Patient Tracker<ul><li>Dedicated navigator registers patient upon referral into a live tracker, which flags patients at risk of missing established milestones</li></ul></li><li>Patient and Provider Support<ul><li>Patient educational brochures, including introduction to care team, commonly used terms and timelines of treatment with contact information for missed milestones or urgent needs</li><li>Patient contact cards, including contact information for neurosurgical and radiation / medical oncology administrative support staff (Sfig 9)</li><li>Program email address, monitored during work hours</li><li>Post-operative IBTP introductory navigator session, with patient support package (SFig 6)</li><li>Navigator follow-up call at 48h, 30 days and 90 days post discharge</li></ul></li><li>Referring physician offsite informational poster (Sfig 8)<ul><li>Referring physician contact cards, including contact information for neurosurgical and radiation / medical oncology administrative support (Sfig 8)</li></ul></li><li>Research and Innovation<ul><li>Standardized introduction to clinical research opportunities and consenting for surgical trials and translational research pre-operatively</li><li>Consenting for epidemiological and cancer therapeutic trials post-operatively during the IBTP introductory educational navigation session</li><li>Gathering clinical epidemiological information prior to discharge, at 30 and 90 days post-op, at progression and at time of death</li></ul></li><li>Operational Efficiency<ul><li>Establishment of key performance indexes and annual accountability metrics</li><li>Annual evaluation of current materials and initiatives through the multi-disciplinary retreat</li><li>Patient feedback forms (Sfig 11)</li></ul></li><li>Outreach and Awareness<ul><li>Creation of program website (SFIb 12)</li><li>Establishment of social media platforms (instagram, facebook (Sfig 13)</li><li>Collaboration with regional and national neuro-oncology outreach events (Sfig – brain tumor week)</li><li>Establishment of “IBPT partners” – local companies who provide reduced rates for housing and food (SFig 10)</li></ul></li></ul> |
| **Test** | Trial the program real time and embed feedback | <ul><li>program launch week, with engagement of regional referring hospitals and official deployment of navigator, live tracker and prototyped materials</li></ul> |

Supplemental Table 1 **IBTP Design Thinking Process Steps** | Stepwise actions taken across each phase of the design thinking model during development of the Integrated Brain Tumour Program. The table outlines key objectives and corresponding actions within the empathize, define, ideate, prototype, and test stages, guiding program design and implementation based on patient and provider input.

| Factor | Analysis |
|---|---|
| **Political** | The IBTP operates within Canada's publicly funded healthcare system, which subjects it to political decision-making, funding models, and provincial health policies. |
| | Regional disparities in rural healthcare are a known political concern, making IBTP politically relevant and potentially well-supported. |
| | Risks include policy discontinuity with changes in provincial government or health authority leadership, which may affect funding and implementation consistency. |
| **Economic** | Budget constraints are a persistent issue in public health, but IBTP may offer cost-saving opportunities by reducing delays, readmissions, and fragmented care. |
| | Economic pressures from an aging population and rising glioblastoma incidence justify the need for scalable, efficient care models. |
| | The program's early-stage, resource-intensive setup presents upfront costs that must be offset by long-term benefits. |
| **Social** | The program directly addresses health equity challenges, particularly rural and Indigenous patient access, making it socially and ethically critical. |
| | There is increasing public and professional awareness around disparities in care, mental health, and culturally competent healthcare. |
| | Demographic factors like age (median GB diagnosis at 64) and rural isolation heighten the social necessity of patient-centered, coordinated programs like IBTP. |
| **Technological** | IBTP integrates digital tools like EMR-linked forms, patient navigation software, and educational materials to streamline care. |
| | Technological literacy, particularly among elderly and rural populations, poses a potential barrier. |
| | There's scope for telehealth expansion and integration with AI tools for triage or predictive analytics in future iterations of IBTP. |
| | The centre transitioned to a new EMR software following IBTP launch which brings substantial provider-education burden amid debugging and patient flow interruption |
| **Legal** | Compliance with provincial privacy regulations (e.g., PHIPA) is essential, especially given the use of EMRs and sensitive patient data for research. |
| | Legal/ethical concerns exist around consent, particularly with vulnerable populations (e.g., cognitive impairment, language barriers, or palliative patients). |
| | Ethical review (already granted) and continuous feedback loops mitigate some of these concerns by embedding patient and provider voice. |
| **Environmental** | Geographic barriers (e.g., remote locations, winter weather) severely impact patient travel, care continuity, and appointment adherence. |
| | Sustainability concerns may emerge with paper-based materials or duplicative assessments, though the move to digital helps. |
| | Future environmental considerations might include green practices in clinical design and telemedicine to reduce travel emissions. |

Supplemental Table 2 **IBTP PESTLE Analysis** | A structured overview of political, economic, social, technological, legal, and environmental influences shaping the design and implementation of the IBTP. This table highlights non-clinical and third-party factors relevant to sustainability, scalability, and real-world integration of a regional brain tumor program.

| Segment | Analysis |
|---|---|
| **Strengths** | ● Human-centered design rooted in empathy and co-design with patients, caregivers, and providers<br>● Interdisciplinary collaboration across neurosurgery, oncology, nursing, administration, and cultural advisors<br>● Mixed-methods evaluation combining quantitative metrics and qualitative insights<br>Strong focus on equity: addresses rural, socioeconomic, and Indigenous disparities directly<br>● Nurse navigator role enhances continuity, emotional support, and care coordination<br>Institutional support and ethics approval provide program legitimacy and infrastructure<br>● Iterative design with real-time feedback loops through a patient working group<br>● Alignment with provincial health quality mandates and learning health system principles |
| **Weaknesses** | ● Resource-intensive: requires dedicated personnel, coordination, and infrastructure<br>● Early-stage: long-term clinical outcomes and scalability are still under evaluation<br>● Role ambiguity: initial lack of clarity around legal documentation and logistical scope for navigators<br>● Sustainability may depend on a few key individuals: risk of burnout or attrition<br>● Fragmented EMR systems hinder seamless communication across departments<br>● Limited generalizability to low-resource or less-connected regions without adaptation |
| **Opportunities** | ● Expansion across Ontario or nationally as a scalable model for other tumor streams<br>● Influence on health policy and funding models for equitable navigation-based care<br>● Stronger integration with psychosocial services, early palliative care, and radiation oncology<br>● Stronger involvement of palliative care through redefining the care model and renaming its function in the IBTP<br>● Enhanced research capacity: early navigator contact supports biobanking and trial recruitment<br>● Development of digital education and symptom tracking tools to extend reach<br>● Positioning as a leader in culturally safe care through the Indigenous navigator model<br>● Knowledge translation via publication, conference presentations, and public engagement |
| **Threats** | ● Policy shifts or funding cuts could deprioritize navigation or innovation programs<br>● Staff burnout due to emotional and administrative load, especially for navigators<br>● Digital inequity: patients with limited access or tech literacy may be left behind<br>● Resistance to cross-departmental integration in siloed care environments<br>● Privacy and legal considerations around digital tools and research documentation<br>● Geographic barriers (e.g., winter travel, limited local resources) persist for rural patients |

Supplemental Table 3 **IBTP SWOT Analysis** | A structured summary of internal strengths and weaknesses, as well as external opportunities and threats, used to guide the early-phase implementation of the IBTP. This analysis informs risk mitigation strategies and highlights potential areas for growth, scalability, and integration into broader neuro-oncology care systems.

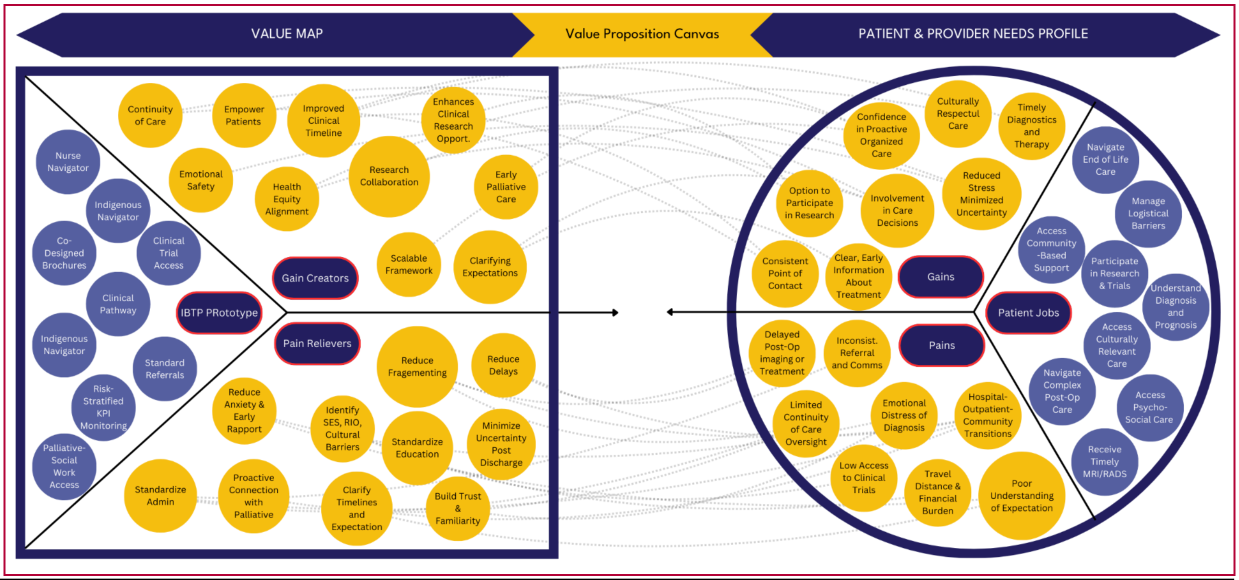

| Responsibilities | Pains | Gains |
|---|---|---|
| • These are the key tasks, needs, or goals the target population is trying to accomplish:<br>• Navigate a complex, fragmented post-operative care system after brain tumor surgery<br>• Receive timely and coordinated access to MRI, radiation, oncology, palliative care, and psychosocial services<br>• Understand their diagnosis, prognosis, and treatment options in a way that is culturally and emotionally accessible<br>• Manage logistical barriers to care (travel, scheduling, follow-ups)<br>• Access community-based support and home care during palliative or end-of-life stages<br>• Participate in research or clinical trials (if desired) | • These are the frustrations, risks, and barriers faced in the existing care system:<br>• Delayed post-op imaging or treatment due to referral or communication breakdowns<br>• Limited continuity and inconsistent provider communication across transitions (hospital → outpatient → community)<br>• Emotional distress from a disjointed or impersonal system, often described as “being dropped off a cliff” after diagnosis<br>• Poor understanding of where to go, what to expect, and who to contact at each step<br>• Travel distance, financial burden, and geographic isolation—especially for rural and marginalized patients<br>• Lack of culturally safe care or tailored education resources<br>• Low access to clinical trials due to urban-centric research infrastructure | • These are the outcomes the patient/caregiver wants or values:<br>• A consistent, compassionate point of contact who knows their story and guides them<br>• Clear, early information about their treatment journey and what to expect<br>• Access to timely diagnostics and therapy that meets provincial benchmarks<br>• Reduced stress through streamlined communication and minimized uncertainty<br>• Culturally respectful care that includes Indigenous or rural navigation<br>• Feeling involved in care decisions and having the option to participate in research<br>• Confidence that their loved one’s care is proactive, organized, and holistic |

| Prototype | Pain Relievers | Gain Creators |
|---|---|---|
| • Nurse navigator role integrated at pre-op, inpatient, and outpatient phases<br>• Indigenous patient navigator to provide culturally grounded, respectful support<br>• Redesigned referral and discharge packages with standardized forms and communication tools<br>• Co-designed educational brochures and digital resources<br>• Navigation-enabled access to palliative care, psychosocial support, and research opportunities<br>• Biobanking and tissue donation coordination<br>• Real-time KPI monitoring with equity-sensitive stratification (e.g., RIO, income quintiles)<br>• Embedded design-thinking methodology to continuously refine interventions | • Reduces fragmentation of care and delays with clear protocols and contact points<br>• Builds trust through early and sustained patient-navigator relationships<br>• Pre-surgical navigator calls alleviate anxiety and establish therapeutic rapport<br>• Identifies socioeconomic, rurality, and cultural barriers early to tailor supports<br>• Proactively connects patients with appropriate community and palliative resources<br>• Clarifies treatment timelines and expectations through standard education and care maps<br>• Minimizes uncertainty post-discharge through scheduled follow-ups and resource access | • Facilitates continuity of care across all phases and settings (hospital, outpatient, community)<br>• Empowers patients and families with knowledge and clear expectations<br>• Improves emotional safety by offering consistent and compassionate contact<br>• Enhances participation in clinical research and informed decision-making<br>• Supports earlier engagement with palliative care, improving quality of life<br>• Aligns care with health equity goals, creating a more just and inclusive model<br>• Builds a scalable, publishable framework that validates design-thinking in neuro-oncology |

Supplemental Table 4 **IBTP Value Proposition Canvas** | Combined visual and tabular summary of the IBTP prototype's value as a minimally viable product. The top diagram maps how the prototype aligns with patient and provider needs, while the bottom table links specific program features to pain relievers and gain creators within the care pathway.

| Patient Case | | |
|---|---|---|
| **Stage** | **Pre-IBTP Experience** | **Post-IBTP (With IBTP Intervention)** |
| **Diagnosis** | ER visit led to rushed surgery; no time for patient education | Navigator connects post-op to explain diagnosis and expected care plan |
| **Post-op Imaging** | MRI delayed >72 hours due to coordination gaps | MRI pre-booked prior to discharge with 48-hour target |
| **Radiation Planning** | Waited 8+ weeks due to referral delays | Referral auto-triggered; radiation consult scheduled within 2–3 weeks |
| **Communication** | No single contact person; confusion about next steps | Patient receives care plan folder and direct phone support from navigator |
| **Travel & Logistics** | Drove 2+ hours multiple times with no coordination | Offered transportation support and coordinated appointments |
| **Follow-up & Support** | Missed psychosocial care; felt alone and overwhelmed | Connected with support groups, community nurse, and Indigenous liaison if relevant |
| **Clinical Trials** | Not informed due to lack of coordination between providers | Navigator screens for eligibility and connects patient to research team |

Supplemental Table 5 **IBTP Patient Case** | A representative patient case highlights key care gaps prior to IBTP and the corresponding interventions introduced post-implementation. Improvements included earlier imaging, streamlined referrals, direct patient communication, transportation support, enhanced psychosocial follow-up, and structured access to clinical trials.

| Provider Case | | |
|---|---|---|
| **Stage** | **Pre-IBTP (Provider Experience)** | **Post-IBTP (With IBTP Intervention)** |
| **Diagnosis** | No standardized workflow for sharing diagnosis; limited patient readiness and poor documentation | Navigator engages immediately post-op; shares diagnostic summary and care map with team and patient |
| **Post-op Imaging** | MRI requisitions often delayed or missed; results not flagged timely | MRI pre-scheduled on EMR prior to discharge; navigator follows up on completion and result distribution |
| **Radiation Planning** | Referrals inconsistently initiated; unclear handoff timing between teams | Standardized referral form triggers automated consult; navigator alerts oncology admin |
| **Communication** | Providers fielded ad hoc calls; unclear points of contact; repeated explanations | Navigator becomes primary contact for logistics, freeing providers for clinical care |
| **Travel & Logistics** | Gaps in understanding patient barriers; missed appointments due to travel or confusion | Navigator consolidates appointments and connects with travel support programs |
| **Follow-up & Support** | Psychosocial needs flagged late or not at all; missed early referral windows | Navigator screens for psychosocial risk, triggers early referrals, and coordinates care team |
| **Clinical Trials** | Providers uncertain of eligibility criteria; missed recruitment opportunities | Navigator pre-screens for eligibility and flags patients in real time via research registry |

Supplemental Table 6 **IBTP Provider Case** | A comparative provider case outlines system-level inefficiencies prior to IBTP and corresponding improvements following implementation. Key enhancements include earlier engagement, standardized referrals, improved documentation, streamlined communication, coordinated logistics, and real-time clinical trial screening, all of which reduced administrative burden and improved care continuity.

#### S5. Analysis-Specific sample sizes and exclusions

| Model | N | Excluded | Events | Non-Event by Timepoint (Censor + Competing Death) |
|---|---|---|---|---|
| **Survival** | 297 | 0 | 162 | 135 |
| **MRI** | 296 | 61 | 234 | 62 |
| **RT** | 296 | 1 | 141 | 155 |
| **Ever-RT** | 200 | 0 | 142 | 58 |

Supplemental Table 7 **End-point Sample Sizes and Exclusions** | Summary of analytic sample sizes, exclusions, and event counts for each primary and secondary endpoint. Sample sizes vary by outcome due to endpoint-specific eligibility criteria, censoring windows, and competing-risk structures. Survival analyses included all eligible patients with no additional exclusions. Timeliness analyses for postoperative MRI and radiotherapy initiation reflect exclusions for missing or non-interpretable timing data. Event counts represent outcome-specific events within the defined analytic windows, while non-events include right-censored observations and, where applicable, competing events such as death.

#### S6. Analysis-Overview

| Analysis | Contrast | HR | LCL | UCL | HR p-value |
|---|---|---|---|---|---|
| **OS (1-year, unadjusted Cox)** | Post vs Pre | 0.581 | 0.368 | 0.916 | 0.021 |
| **OS (14-day landmark, unadjusted Cox)** | Post vs Pre | 0.604 | 0.378 | 0.964 | 0.034 |
| **OS (14-day landmark, covar adjusted)** | Post vs Pre | 0.62 | 0.38 | 0.99 | 0.046 |
| **OS (90-Day, unadjusted)** | Post vs Pre | 0.62 | 0.31 | 1.25 | 0.239 |
| **Landmark death sensitivity - dod −30** | Post vs Pre | 0.598 | 0.368 | 0.971 | 0.038 |
| **Landmark death sensitivity - dod +30** | Post vs Pre | 0.595 | 0.378 | 0.937 | 0.025 |
| **WHO-2021 subset (≥2021 surgeries, landmark)** | Post vs Pre | 0.571 | 0.351 | 0.929 | 0.024 |
| **OS ITS Adjusted - HR at 0 months** | Post vs counterfactual | 1.236 | 0.46 | 3.322 | NA |
| **OS ITS Adjusted - HR at 6 months** | Post vs counterfactual | 0.438 | 0.102 | 1.876 | NA |
| **OS ITS Adjusted - HR at 12 months** | Post vs counterfactual | 0.155 | 0.016 | 1.498 | NA |
| **OS ITS Adjusted Calendar Spline** | Post vs counterfactual | 0.11 | NA | NA | 0.002 |
| **OS (1-Year, Calendar Adjusted)** | Post vs Pre | 0.94 | 0.29 | 3.29 | 0.93 |
| **OS (Bootstrap percentile CI)** | Post vs Pre | 0.604 | 0.369 | 0.924 | NA |
| **MRI (48-Hour, unadjusted Cox)** | Post vs Pre | 1.76 | 1.20 | 2.58 | 0.021 |
| **MRI (7-day, unadjusted Cox)** | Post vs Pre | 1.78 | 1.24 | 2.55 | 0.024 |
| **MRI (48-Hour, covar adjusted)** | Post vs Pre | 1.56 | 1.07 | 2.28 | 0.021 |
| **MRI (48-Hour, unadjusted Fine-Gray)** | Post vs Pre | 1.61 | 1.21 | 2.12 | 0.020 |
| **MRI (48-Hour, covar adjusted Fine-Gray)** | Post vs Pre | 1.42 | 1.06 | 1.89 | 0.019 |
| **MRI (Bootstrap percentile CI)** | Post vs Pre | 1.50 | 1.03 | 2.21 | NA |
| **RT (6-week, unadjusted Cox)** | Post vs Pre | 1.12 | 0.71 | 1.78 | 0.618 |
| **RT (6-week, covar adjusted)** | Post vs Pre | 1.03 | 0.64 | 1.68 | 0.89 |
| **RT (6-week, unadjusted Fine-Gray)** | Post vs Pre | 0.94 | 0.59 | 1.48 | 0.783 |
| **RT (6-week, covar adjusted Fine-Gray)** | Post vs Pre | 1.04 | 0.66 | 1.65 | 0.865 |
| **RT (Bootstrap percentile CI)** | Post vs Pre | 1.07 | 0.67 | 1.64 | NA |
| **Survival Rurality Interaction (1-Year, Pre)** | Rural vs Urban | 1.369 | 0.979 | 1.913 | 0.066 |
| **Survival Rurality Interaction (1-Year, Post)** | Rural vs Urban | 1.338 | 0.573 | 3.124 | 0.501 |
| **Survival Rurality Interaction Term 1-Year** | Post vs Pre | 0.978 | 0.393 | 2.432 | 0.961 |
| **MRI Rurality Interaction (48-Hour, Pre)** | Rural vs Urban | 0.883 | 0.681 | 1.145 | 0.349 |
| **MRI Rurality Interaction (48-Hour, Post)** | Rural vs Urban | 0.727 | 0.410 | 1.287 | 0.274 |
| **MRI Rurality Interaction Term** | Post vs Pre | 0.823 | 0.440 | 1.540 | 0.542 |
| **RT Rurality Interaction (6-Week, Pre)** | Rural vs Urban | 0.595 | 0.416 | 0.850 | 0.004 |

| | | | | | |
|---|---|---|---|---|---|
| **RT Rurality Interaction (6-Week, Posts)** | Rural vs Urban | 1.963 | 0.827 | 4.659 | 0.126 |
| **RT Rurality Interaction Term** | Post vs Pre | 3.301 | 1.295 | 8.413 | 0.012 |

Supplemental Table 8 **Model Estimates Across Sensitivity Analyses** | Hazard ratio estimates across primary analyses, sensitivity analyses, calendar-time–adjusted models, competing-risk models, bootstrap resampling, and rurality interaction analyses. Estimates are reported as hazard ratios (HRs) with 95% confidence intervals (CIs) and corresponding p-values where applicable. Models include unadjusted and covariate-adjusted Cox proportional hazards models, Fine–Gray competing-risk models for timeliness outcomes, interrupted time series (ITS) models, and landmark analyses. Bootstrap confidence intervals were derived using percentile methods. Interaction terms assess differential effects by rurality status within pre- and post-implementation periods.

**S7. Covariate Adjusted Model Effects**

| Covariate | HR Effect | LCL | UCL | P-Value |
|---|---|---|---|---|
| **Cohort** | 0.62 | 0.38 | 0.99 | 0.046 |
| **Age (10-Year)** | 1.56 | 1.29 | 1.88 | <0.01 |
| **Sex** | 0.91 | 0.64 | 1.28 | 0.59 |
| **Rurality** | 1.14 | 0.80 | 1.61 | 0.47 |
| **MGMT** | 0.91 | 0.65 | 1.27 | 0.58 |

Supplemental Table 8 **Covariate-Adjusted One-Year Overall Survival** | Adjusted hazard ratios (HRs) and 95% confidence intervals from a multivariable Cox proportional hazards model evaluating one-year overall survival. The model includes cohort (Post-IBTP vs Pre-IBTP), age (per 10-year increase), sex, rurality, and MGMT promoter methylation status (excluding unknown). Proportional hazards assumptions were satisfied for all included covariates.

| Covariate | HR Effect | LCL | UCL | P-Value |
|---|---|---|---|---|
| **Cohort** | 1.56 | 1.07 | 2.28 | 0.021 |
| **Age (10-Year)** | 1.01 | 0.88 | 1.16 | 0.89 |
| **Sex** | 0.87 | 0.63 | 1.22 | 0.43 |
| **Rurality** | 0.79 | 0.57 | 1.10 | 0.16 |
| **MGMT** | 0.89 | 0.63 | 1.24 | 0.49 |

Supplemental Table 9 **Covariate-Adjusted Timeliness of Postoperative MRI** | Adjusted hazard ratios (HRs) and 95% confidence intervals from a multivariable Cox proportional hazards model evaluating receipt of postoperative MRI within 48 hours of surgery. The model includes cohort (Post-IBTP vs Pre-IBTP), age (per 10-year increase), sex, rurality, and MGMT promoter methylation status (excluding unknown). After model refinement, proportional hazards assumptions were satisfied for all covariates except cohort; complementary Fine–Gray competing-risk and bootstrap sensitivity analyses were performed to support inference.

| Covariate | HR Effect | LCL | UCL | P-Value |
|---|---|---|---|---|
| **Cohort** | 0.99 | 0.62 | 1.57 | 0.96 |
| **Age (10-Year)** | 0.87 | 0.76 | 1.01 | 0.07 |
| **Sex** | 1.08 | 0.77 | 1.51 | 0.66 |
| **Rurality** | 0.77 | 0.55 | 1.09 | 0.14 |
| **MGMT** | 1.15 | 0.82 | 1.62 | 0.42 |

Supplemental Table 10 **Covariate-Adjusted Timeliness of Postoperative Radiotherapy** | Adjusted hazard ratios (HRs) and 95% confidence intervals from a multivariable Cox proportional hazards model evaluating initiation of radiotherapy within six weeks of surgery. The model includes cohort (Post-IBTP vs Pre-IBTP), age (per 10-year increase), sex, rurality, and MGMT promoter methylation status (excluding unknown). Proportional hazards assumptions were satisfied for all included covariates.

##### S9. Proportional Hazard Assumptions Testing

| Covariate | Measurement | PH Test p-Value | Violation Handled? |
|---|---|---|---|
| **Cohort** | Baseline | 0.369 | No |
| **Age (10-Year)** | Baseline | 0.891 | No |
| **Sex** | Baseline | 0.958 | No |
| **Rurality** | Baseline | 0.917 | No |

Supplemental Table 11 **Survival COX Proportional Hazard Covariate Assumptions** | Results of proportional hazards (PH) assumption testing for covariates included in Cox proportional hazards models evaluating one-year overall survival. PH assumptions were assessed using Schoenfeld residual–based tests. No violations were detected for cohort, age, sex, or rurality, and no corrective modeling strategies were required.

| Covariate | Measurement | PH Test p-Value | Violation Handled? |
|---|---|---|---|
| **Cohort** | Baseline | 0.033 | Fine-Gray |
| **Age (10-Year)** | Baseline | 0.734 | No |
| **Sex** | Baseline | 0.926 | No |
| **Rurality** | Baseline | 0.191 | No |

Supplemental Table 12 **Postoperative MRI COX Proportional Hazard Covariate Assumptions** | Results of proportional hazards (PH) assumption testing for covariates included in Cox proportional hazards models evaluating postoperative MRI within 48 hours of surgery. A violation of the PH assumption was detected for cohort status. This was addressed through complementary Fine–Gray competing-risk models and bootstrap sensitivity analyses, while other covariates satisfied PH assumptions without adjustment.

| Covariate | Measurement | PH Test p-Value | Violation Handled? |
|---|---|---|---|
| **Cohort** | Baseline | 0.191 | No |
| **Age (10-Year)** | Baseline | 0.324 | No |
| **Sex** | Baseline | 0.341 | No |
| **Rurality** | Baseline | 0.443 | No |

Supplemental Table 13 **Postoperative Radiotherapy COX Proportional Hazard Covariate Assumptions** | Results of proportional hazards (PH) assumption testing for covariates included in Cox proportional hazards models evaluating initiation of radiotherapy within six weeks of surgery. No violations of the proportional hazards assumption were detected for cohort, age, sex, or rurality.

#### Supplementary Figures

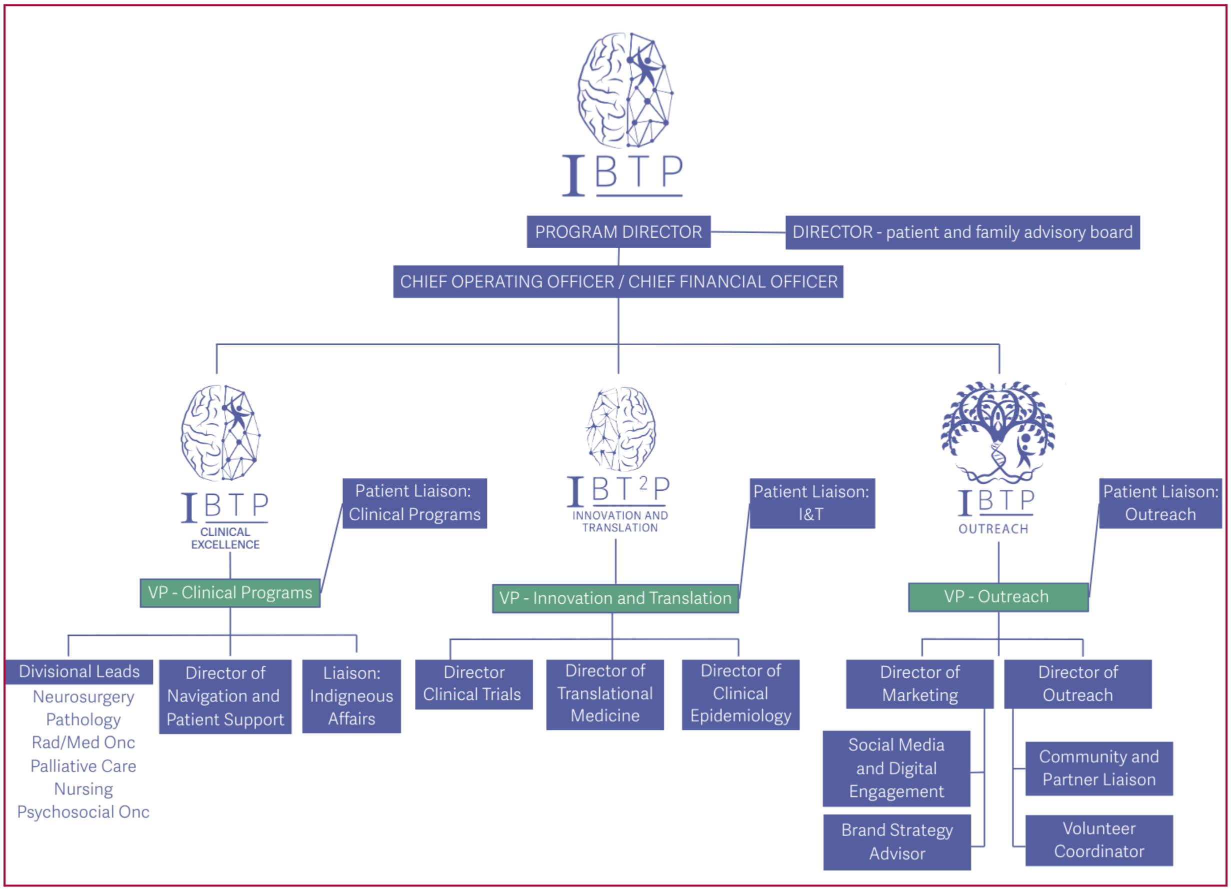

Supplemental Figure 2 **Organizational Structure of the Integrated Brain Tumour Program |** This diagram illustrates the governance model of the IBTP, which is organized into three core divisions: Clinical (ICEiN), Innovation and Translation (IBT2P), and Outreach. Each division includes operational leads from neurosurgery, oncology, navigation, research, and education. The structure promotes integrated leadership and coordination across clinical care, research, and stakeholder engagement. Green boxes represent positions opening in the next iteration of the IBTP, while blue boxes represent positions deployed during the initial launch.

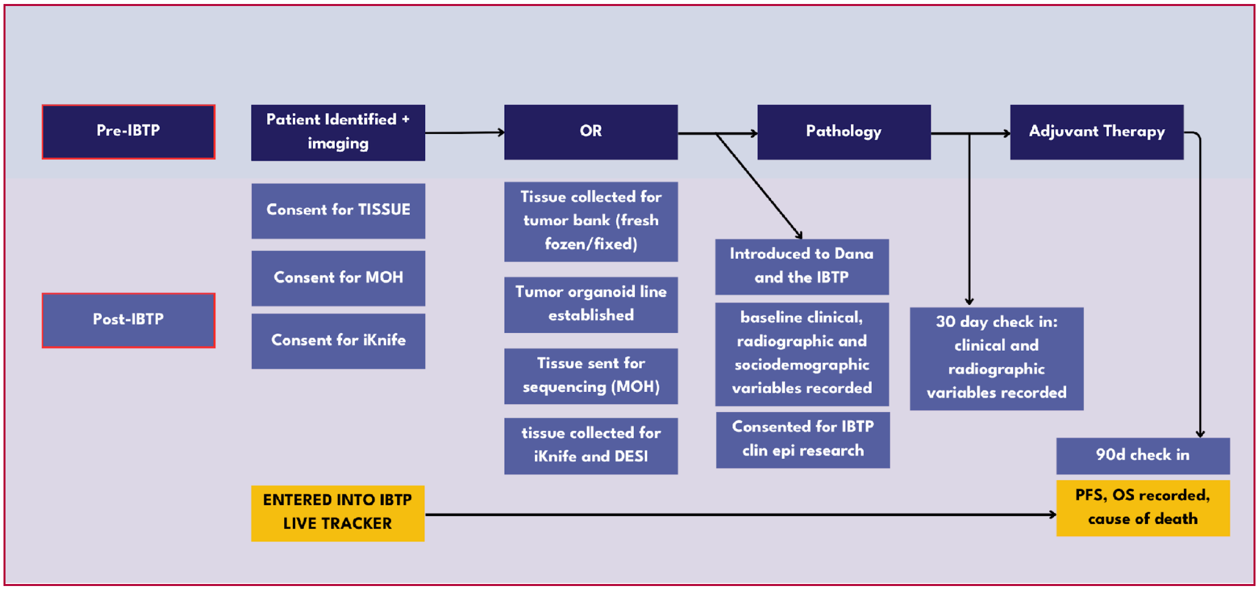

Supplemental Figure 3A **IBTP pathway integration into standard clinical care** | Schematic showing the patient pathway before and after IBTP implementation. The top sequence represents the pre-IBTP clinical flow. The lower steps illustrate post-IBTP additions including research consent, intraoperative tissue protocols, onboarding into the IBTP live tracker, and structured check-ins at 30 and 90 days. These changes support early engagement, enhanced sample collection, and consistent longitudinal follow-up.

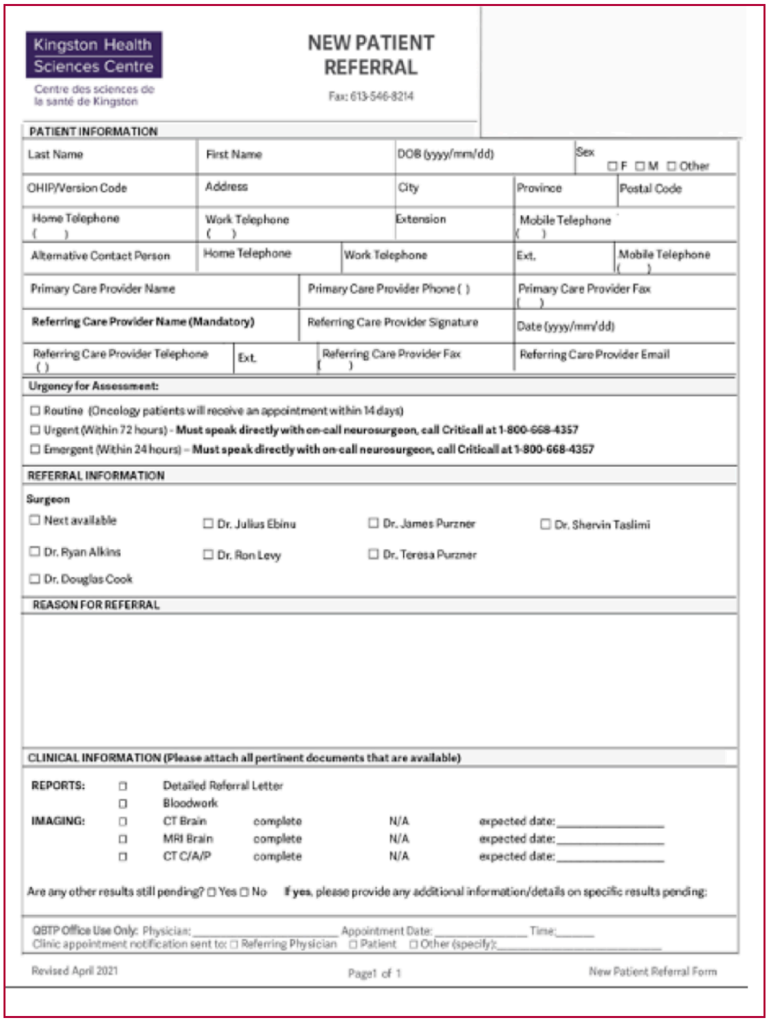

Kingston Health Sciences Centre
Centre des sciences de la santé de Kingston

**NEW PATIENT REFERRAL**

PATIENT INFORMATION

| Last Name | First Name | DOB (yyyy/mm/dd) | Sex ☐ F ☐ M ☐ Other |
|---|---|---|---|
| OHIP/Version Code | Address | City | Province | Postal Code |
| Home Telephone ( ) | Work Telephone ( ) | Extension | Mobile Telephone ( ) |
| Alternative Contact Person | Home Telephone | Work Telephone | Ext. | Mobile Telephone ( ) |
| Primary Care Provider Name | Primary Care Provider Phone ( ) | Primary Care Provider Fax ( ) |
| **Referring Care Provider Name (Mandatory)** | Referring Care Provider Signature | Date (yyyy/mm/dd) |
| Referring Care Provider Telephone ( ) | Ext. | Referring Care Provider Fax ( ) | Referring Care Provider Email |

**Urgency for Assessment:**

☐ Routine (Oncology patients will receive an appointment within 14 days)
☐ Urgent (Within 72 hours) - **Must speak directly with on-call neurosurgeon, call Criticall at 1-800-668-4357**
☐ Emergent (Within 24 hours) – **Must speak directly with on-call neurosurgeon, call Criticall at 1-800-668-4357**

REFERRAL INFORMATION

**Surgeon**

☐ Next available ☐ Dr. Julius Ebinu ☐ Dr. James Purzner ☐ Dr. Shervin Taslimi
☐ Dr. Ryan Alkins ☐ Dr. Ron Levy ☐ Dr. Teresa Purzner
☐ Dr. Douglas Cook

REASON FOR REFERRAL

CLINICAL INFORMATION **(Please attach all pertinent documents that are available)**

**REPORTS:** ☐ Detailed Referral Letter
☐ Bloodwork
**IMAGING:** ☐ CT Brain complete N/A expected date: ____________
☐ MRI Brain complete N/A expected date: ____________
☐ CT C/A/P complete N/A expected date: ____________

Are any other results still pending? ☐ Yes ☐ No **If yes**, please provide any additional information/details on specific results pending:

QBTP Office Use Only: Physician: ____________ Appointment Date: ____________ Time: ______
Clinic appointment notification sent to: ☐ Referring Physician ☐ Patient ☐ Other (specify): ____________

Revised April 2021 Page1 of 1 New Patient Referral Form

Supplemental Figure 4 **Standardized referral form for the Integrated Brain Tumour Program (IBTP)** | Referral form used to initiate neurosurgical consultation for patients with suspected or newly diagnosed glioblastoma. The form captures demographic details, referring provider information, urgency of referral, and relevant imaging or laboratory data. It also allows for preferred surgeon designation to streamline triage and surgical planning.

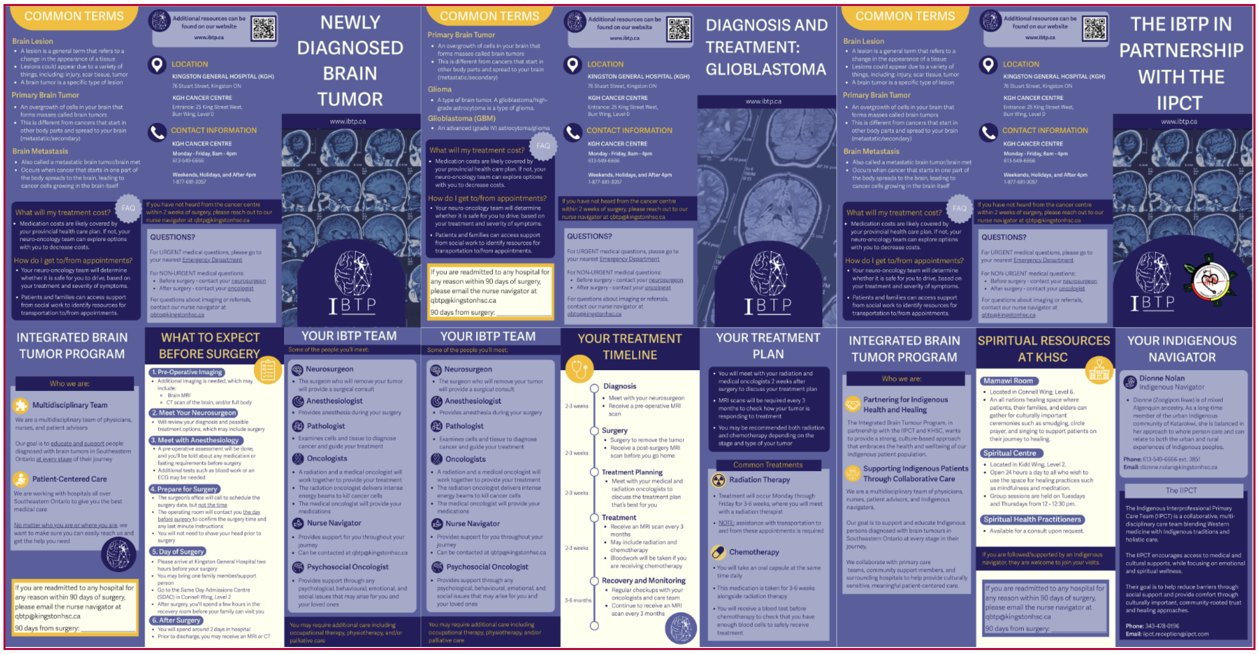

Supplemental Figure 5 **IBTP Patient Education Brochures** | Tri-fold patient education brochures designed to support patients newly diagnosed with brain tumours. These brochures include contact information, timelines for care, frequently asked questions, and key definitions to reduce confusion, clarify next steps, and enhance perioperative preparedness. The timeline-based layout and visual emphasis on navigation aim to reduce loss to follow-up and improve early engagement.

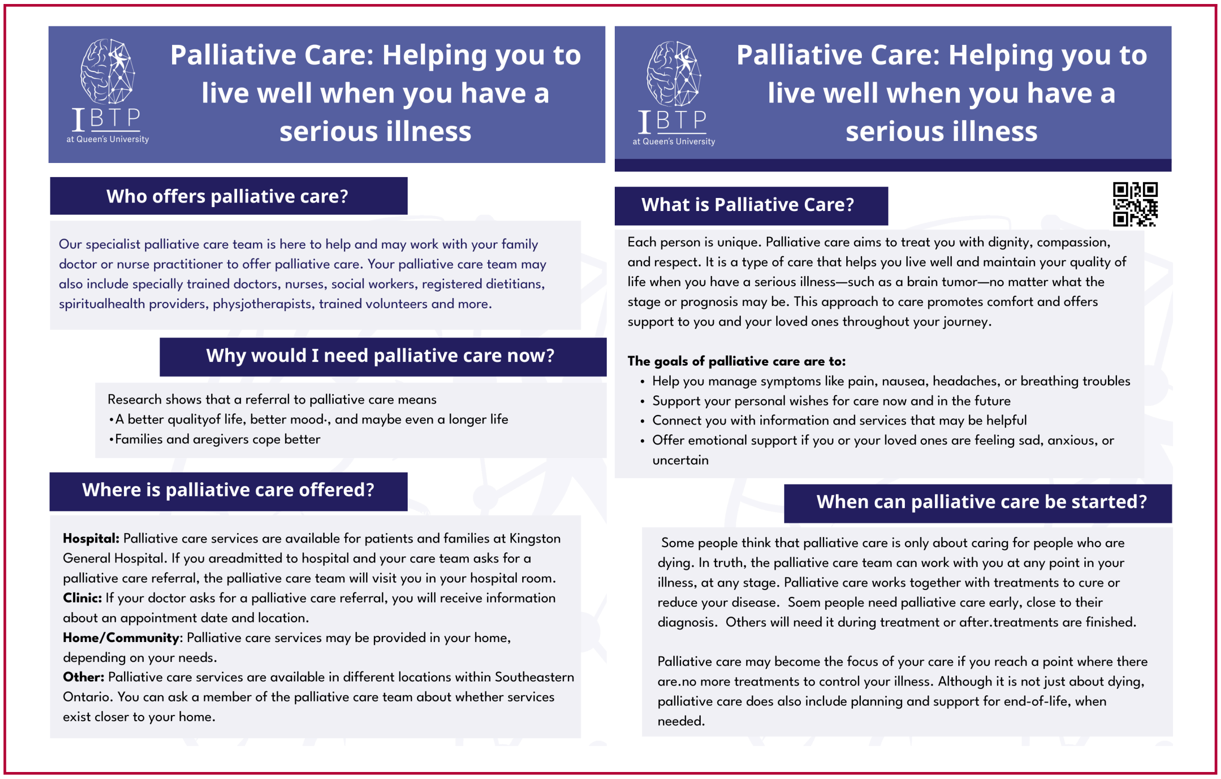

#### Palliative Care: Helping you to live well when you have a serious illness

IBTP at Queen's University

##### Who offers palliative care?

Our specialist palliative care team is here to help and may work with your family doctor or nurse practitioner to offer palliative care. Your palliative care team may also include specially trained doctors, nurses, social workers, registered dietitians, spiritualhealth providers, physjotherapists, trained volunteers and more.

##### Why would I need palliative care now?

Research shows that a referral to palliative care means
•A better qualityof life, better mood·, and maybe even a longer life
•Families and aregivers cope better

##### Where is palliative care offered?

**Hospital:** Palliative care services are available for patients and families at Kingston General Hospital. If you areadmitted to hospital and your care team asks for a palliative care referral, the palliative care team will visit you in your hospital room.
**Clinic:** If your doctor asks for a palliative care referral, you will receive information about an appointment date and location.
**Home/Community**: Palliative care services may be provided in your home, depending on your needs.
**Other:** Palliative care services are available in different locations within Southeastern Ontario. You can ask a member of the palliative care team about whether services exist closer to your home.

#### Palliative Care: Helping you to live well when you have a serious illness

IBTP at Queen's University

##### What is Palliative Care?

Each person is unique. Palliative care aims to treat you with dignity, compassion, and respect. It is a type of care that helps you live well and maintain your quality of life when you have a serious illness—such as a brain tumor—no matter what the stage or prognosis may be. This approach to care promotes comfort and offers support to you and your loved ones throughout your journey.

**The goals of palliative care are to:**

- Help you manage symptoms like pain, nausea, headaches, or breathing troubles
- Support your personal wishes for care now and in the future
- Connect you with information and services that may be helpful
- Offer emotional support if you or your loved ones are feeling sad, anxious, or uncertain

##### When can palliative care be started?

Some people think that palliative care is only about caring for people who are dying. In truth, the palliative care team can work with you at any point in your illness, at any stage. Palliative care works together with treatments to cure or reduce your disease. Soem people need palliative care early, close to their diagnosis. Others will need it during treatment or after.treatments are finished.

Palliative care may become the focus of your care if you reach a point where there are.no more treatments to control your illness. Although it is not just about dying, palliative care does also include planning and support for end-of-life, when needed.

Supplemental Figure 6 **IBTP Palliative Care Education Forms** | Patient-facing palliative care education materials provided at the initial visit. These documents address common misconceptions, outline the benefits and timing of palliative care, and describe where services are available. The content is designed to reduce fear, normalize early referral, and support informed discussions about goals of care throughout the patient's treatment journey.

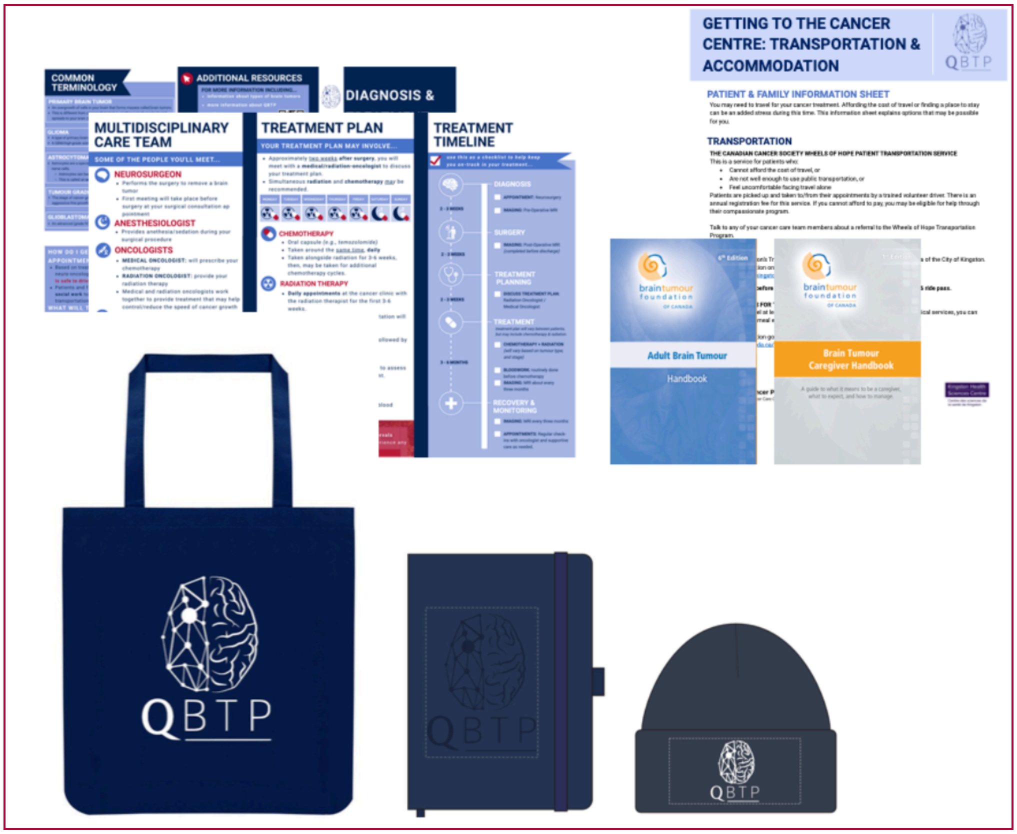

Supplemental Figure 7 **IBTP Onboarding Package** | The IBTP onboarding package includes a branded bag containing educational materials, appointment documentation tools, and patient comfort items. The package is provided to patients once they are entered into the IBTP live tracker. It includes a notebook for recording clinical visits, a hat for patients experiencing hair loss due to surgery or chemotherapy, and patient information brochures. Also included are Brain Tumour Foundation of Canada resources, parking and travel details, and clinical pathway materials designed to ensure that each patient has a clear and coordinated care plan throughout neurosurgical management.

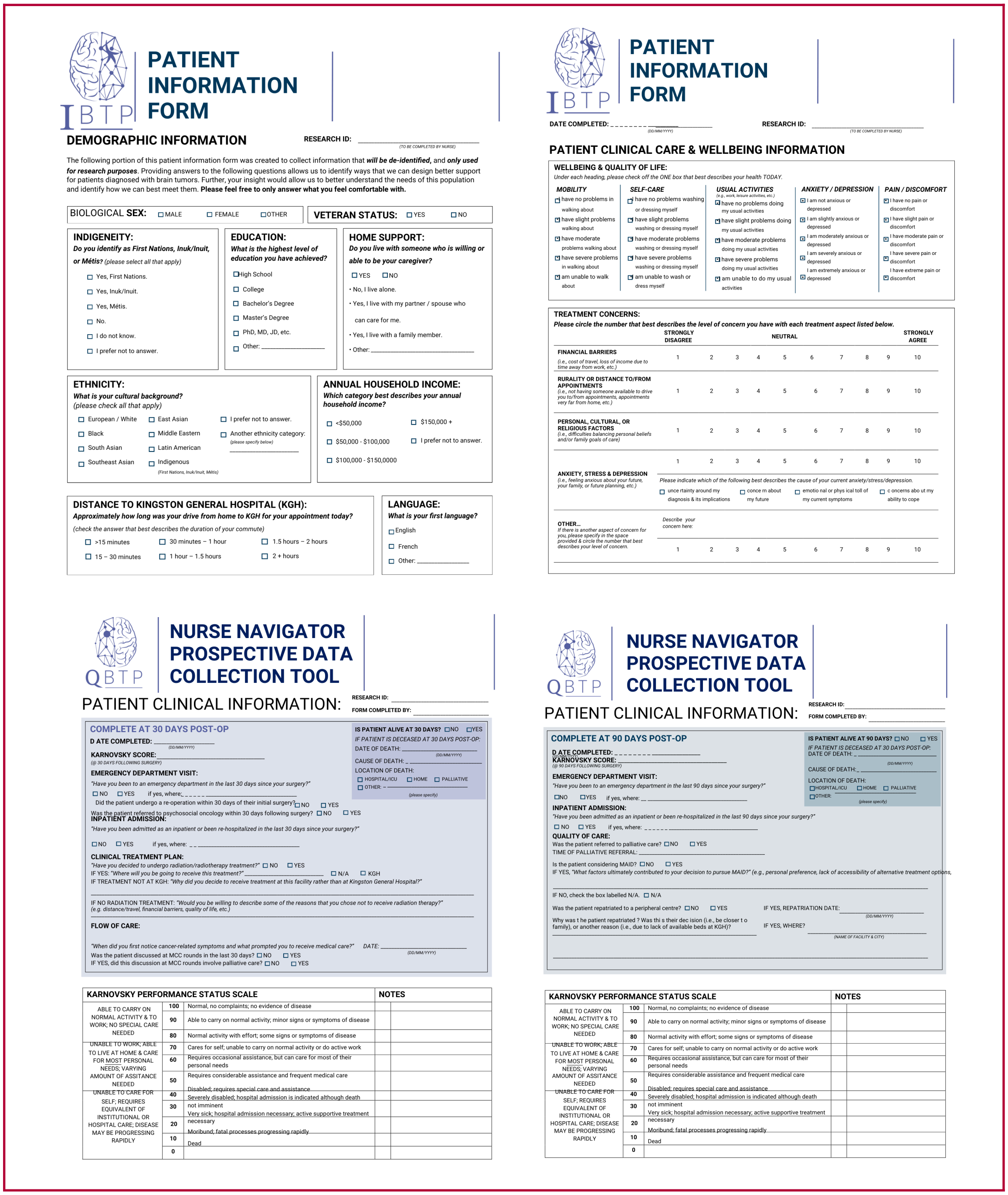

### PATIENT INFORMATION FORM

#### DEMOGRAPHIC INFORMATION

RESEARCH ID: ____________ (TO BE COMPLETED BY NURSE)

The following portion of this patient information form was created to collect information that ***will be de-identified***, and ***only used for research purposes***. Providing answers to the following questions allows us to identify ways that we can design better support for patients diagnosed with brain tumors. Further, your insight would allow us to better understand the needs of this population and identify how we can best meet them. **Please feel free to only answer what you feel comfortable with.**

BIOLOGICAL **SEX:** ☐ MALE ☐ FEMALE ☐ OTHER

**VETERAN STATUS:** ☐ YES ☐ NO

**INDIGENEITY:**
***Do you identify as First Nations, Inuk/Inuit, or Métis?*** *(please select all that apply)*
- ☐ Yes, First Nations.
- ☐ Yes, Inuk/Inuit.
- ☐ Yes, Métis.
- ☐ No.
- ☐ I do not know.
- ☐ I prefer not to answer.

**EDUCATION:**
***What is the highest level of education you have achieved?***
- ☐ High School
- ☐ College
- ☐ Bachelor's Degree
- ☐ Master's Degree
- ☐ PhD, MD, JD, etc.
- ☐ Other: ____________

**HOME SUPPORT:**
***Do you live with someone who is willing or able to be your caregiver?***
☐ YES ☐ NO
- No, I live alone.
- Yes, I live with my partner / spouse who can care for me.
- Yes, I live with a family member.
- Other: ____________

**ETHNICITY:**
***What is your cultural background?***
*(please check all that apply)*
- ☐ European / White
- ☐ Black
- ☐ South Asian
- ☐ Southeast Asian
- ☐ East Asian
- ☐ Middle Eastern
- ☐ Latin American
- ☐ Indigenous *(First Nations, Inuk/Inuit, Métis)*
- ☐ I prefer not to answer.
- ☐ Another ethnicity category: *(please specify below)* ____________

**ANNUAL HOUSEHOLD INCOME:**
***Which category best describes your annual household income?***
- ☐ <$50,000
- ☐ $50,000 - $100,000
- ☐ $100,000 - $150,0000
- ☐ $150,000 +
- ☐ I prefer not to answer.

**DISTANCE TO KINGSTON GENERAL HOSPITAL (KGH):**
***Approximately how long was your drive from home to KGH for your appointment today?***
*(check the answer that best describes the duration of your commute)*
- ☐ >15 minutes
- ☐ 15 – 30 minutes
- ☐ 30 minutes – 1 hour
- ☐ 1 hour – 1.5 hours
- ☐ 1.5 hours – 2 hours
- ☐ 2 + hours

**LANGUAGE:**
***What is your first language?***
- ☐ English
- ☐ French
- ☐ Other: ____________

### PATIENT INFORMATION FORM

DATE COMPLETED: ____________ (DD/MM/YYYY) RESEARCH ID: ____________ (TO BE COMPLETED BY NURSE)

#### PATIENT CLINICAL CARE & WELLBEING INFORMATION

**WELLBEING & QUALITY OF LIFE:**
*Under each heading, please check off the ONE box that best describes your health TODAY.*

***MOBILITY***
- ☐ I have no problems in walking about
- ☐ I have slight problems walking about
- ☐ I have moderate problems walking about
- ☐ I have severe problems in walking about
- ☐ am unable to walk about

***SELF-CARE***
- ☐ I have no problems washing or dressing myself
- ☐ I have slight problems washing or dressing myself
- ☐ I have moderate problems washing or dressing myself
- ☐ I have severe problems washing or dressing myself
- ☐ am unable to wash or dress myself

***USUAL ACTIVITIES*** *(e.g., work, leisure activities, etc.)*
- ☐ I have no problems doing my usual activities
- ☐ I have slight problems doing my usual activities
- ☐ I have moderate problems doing my usual activities
- ☐ I have severe problems doing my usual activities
- ☐ am unable to do my usual activities

***ANXIETY / DEPRESSION***
- ☐ I am not anxious or depressed
- ☐ I am slightly anxious or depressed
- ☐ I am moderately anxious or depressed
- ☐ I am severely anxious or depressed
- ☐ I am extremely anxious or depressed

***PAIN / DISCOMFORT***
- ☐ I have no pain or discomfort
- ☐ I have slight pain or discomfort
- ☐ I have moderate pain or discomfort
- ☐ I have severe pain or discomfort
- ☐ I have extreme pain or discomfort

**TREATMENT CONCERNS:**
***Please circle the number that best describes the level of concern you have with each treatment aspect listed below.***

| | STRONGLY DISAGREE | | | | NEUTRAL | | | | | STRONGLY AGREE |
|---|---|---|---|---|---|---|---|---|---|---|
| **FINANCIAL BARRIERS** *(i.e., cost of travel, loss of income due to time away from work, etc.)* | 1 | 2 | 3 | 4 | 5 | 6 | 7 | 8 | 9 | 10 |
| **RURALITY OR DISTANCE TO/FROM APPOINTMENTS** *(i.e., not having someone available to drive you to/from appointments, appointments very far from home, etc.)* | 1 | 2 | 3 | 4 | 5 | 6 | 7 | 8 | 9 | 10 |
| **PERSONAL, CULTURAL, OR RELIGIOUS FACTORS** *(i.e., difficulties balancing personal beliefs and/or family goals of care)* | 1 | 2 | 3 | 4 | 5 | 6 | 7 | 8 | 9 | 10 |
| **ANXIETY, STRESS & DEPRESSION** *(i.e., feeling anxious about your future, your family, or future planning, etc.)* | 1 | 2 | 3 | 4 | 5 | 6 | 7 | 8 | 9 | 10 |
| **OTHER…** *If there is another aspect of concern for you, please specify in the space provided & circle the number that best describes your level of concern.* | 1 | 2 | 3 | 4 | 5 | 6 | 7 | 8 | 9 | 10 |

*Please indicate which of the following best describes the cause of your current anxiety/stress/depression.*
☐ unce rtainty around my diagnosis & its implications ☐ conce rn about my future ☐ emotio nal or phys ical toll of my current symptoms ☐ c oncerns abo ut my ability to cope

*Describe your concern here:*

### NURSE NAVIGATOR PROSPECTIVE DATA COLLECTION TOOL

#### PATIENT CLINICAL INFORMATION:

RESEARCH ID: ____________
FORM COMPLETED BY: ____________

##### COMPLETE AT 30 DAYS POST-OP

**D ATE COMPLETED:** ____________ (DD/MM/YYYY)

**KARNOVSKY SCORE:** ____________ (@ 30 DAYS FOLLOWING SURGERY)

**IS PATIENT ALIVE AT 30 DAYS?** ☐ NO ☐ YES
*IF PATIENT IS DECEASED AT 30 DAYS POST-OP:*
DATE OF DEATH: ____________ (DD/MM/YYYY)
CAUSE OF DEATH: ____________
LOCATION OF DEATH: ☐ HOSPITAL/ICU ☐ HOME ☐ PALLIATIVE ☐ OTHER: ____________ *(please specify)*

**EMERGENCY DEPARTMENT VISIT:**
*"Have you been to an emergency department in the last 30 days since your surgery?"*
☐ NO ☐ YES if yes, where: ____________
Did the patient undergo a re-operation within 30 days of their initial surgery? ☐ NO ☐ YES
Was the patient referred to psychosocial oncology within 30 days following surgery? ☐ NO ☐ YES

**INPATIENT ADMISSION:**
*"Have you been admitted as an inpatient or been re-hospitalized in the last 30 days since your surgery?"*
☐ NO ☐ YES if yes, where: ____________

**CLINICAL TREATMENT PLAN:**
*"Have you decided to undergo radiation/radiotherapy treatment?"* ☐ NO ☐ YES
IF YES: *"Where will you be going to receive this treatment?"* ____________ ☐ N/A ☐ KGH
IF TREATMENT NOT AT KGH: *"Why did you decide to receive treatment at this facility rather than at Kingston General Hospital?"*
IF NO RADIATION TREATMENT: *"Would you be willing to describe some of the reasons that you chose not to receive radiation therapy?" (e.g. distance/travel, financial barriers, quality of life, etc.)*

**FLOW OF CARE:**
*"When did you first notice cancer-related symptoms and what prompted you to receive medical care?"* DATE: ____________ (DD/MM/YYYY)
Was the patient discussed at MCC rounds in the last 30 days? ☐ NO ☐ YES
IF YES, did this discussion at MCC rounds involve palliative care? ☐ NO ☐ YES

| KARNOVSKY PERFORMANCE STATUS SCALE | | | NOTES |
|---|---|---|---|
| ABLE TO CARRY ON NORMAL ACTIVITY & TO WORK; NO SPECIAL CARE NEEDED | 100 | Normal, no complaints; no evidence of disease | |
| | 90 | Able to carry on normal activity; minor signs or symptoms of disease | |
| | 80 | Normal activity with effort; some signs or symptoms of disease | |
| UNABLE TO WORK; ABLE TO LIVE AT HOME & CARE FOR MOST PERSONAL NEEDS; VARYING AMOUNT OF ASSITANCE NEEDED | 70 | Cares for self; unable to carry on normal activity or do active work | |
| | 60 | Requires occasional assistance, but can care for most of their personal needs | |
| | 50 | Requires considerable assistance and frequent medical care | |
| UNABLE TO CARE FOR SELF; REQUIRES EQUIVALENT OF INSTITUTIONAL OR HOSPITAL CARE; DISEASE MAY BE PROGRESSING RAPIDLY | 40 | Disabled; requires special care and assistance | |
| | 30 | Severely disabled; hospital admission is indicated although death not imminent | |
| | 20 | Very sick; hospital admission necessary; active supportive treatment necessary | |
| | 10 | Moribund; fatal processes progressing rapidly | |
| | 0 | Dead | |

### NURSE NAVIGATOR PROSPECTIVE DATA COLLECTION TOOL

#### PATIENT CLINICAL INFORMATION:

RESEARCH ID: ____________
FORM COMPLETED BY: ____________

##### COMPLETE AT 90 DAYS POST-OP

**D ATE COMPLETED:** ____________ (DD/MM/YYYY)

**KARNOVSKY SCORE:** ____________ (@ 90 DAYS FOLLOWING SURGERY)

**IS PATIENT ALIVE AT 90 DAYS?** ☐ NO ☐ YES
*IF PATIENT IS DECEASED AT 30 DAYS POST-OP:*
DATE OF DEATH: ____________ (DD/MM/YYYY)
CAUSE OF DEATH: ____________
LOCATION OF DEATH: ☐ HOSPITAL/ICU ☐ HOME ☐ PALLIATIVE ☐ OTHER: ____________ *(please specify)*

**EMERGENCY DEPARTMENT VISIT:**
*"Have you been to an emergency department in the last 90 days since your surgery?"*
☐ NO ☐ YES if yes, where: ____________

**INPATIENT ADMISSION:**
*"Have you been admitted as an inpatient or been re-hospitalized in the last 90 days since your surgery?"*
☐ NO ☐ YES if yes, where: ____________

**QUALITY OF CARE:**
Was the patient referred to palliative care? ☐ NO ☐ YES
TIME OF PALLIATIVE REFERRAL: ____________

Is the patient considering MAID? ☐ NO ☐ YES
IF YES, *"What factors ultimately contributed to your decision to pursue MAID?" (e.g., personal preference, lack of accessibility of alternative treatment options,*

IF NO, check the box labelled N/A. ☐ N/A

Was the patient repatriated to a peripheral centre? ☐ NO ☐ YES IF YES, REPATRIATION DATE: ____________ (DD/MM/YYYY)

Why was t he patient repatriated ? Was thi s their dec ision (i.e., be closer t o family), or another reason (i.e., due to lack of available beds at KGH)?

IF YES, WHERE? ____________ (NAME OF FACILITY & CITY)

| KARNOVSKY PERFORMANCE STATUS SCALE | | | NOTES |
|---|---|---|---|
| ABLE TO CARRY ON NORMAL ACTIVITY & TO WORK; NO SPECIAL CARE NEEDED | 100 | Normal, no complaints; no evidence of disease | |
| | 90 | Able to carry on normal activity; minor signs or symptoms of disease | |
| | 80 | Normal activity with effort; some signs or symptoms of disease | |
| UNABLE TO WORK; ABLE TO LIVE AT HOME & CARE FOR MOST PERSONAL NEEDS; VARYING AMOUNT OF ASSITANCE NEEDED | 70 | Cares for self; unable to carry on normal activity or do active work | |
| | 60 | Requires occasional assistance, but can care for most of their personal needs | |
| | 50 | Requires considerable assistance and frequent medical care | |
| UNABLE TO CARE FOR SELF; REQUIRES EQUIVALENT OF INSTITUTIONAL OR HOSPITAL CARE; DISEASE MAY BE PROGRESSING RAPIDLY | 40 | Disabled; requires special care and assistance | |
| | 30 | Severely disabled; hospital admission is indicated although death not imminent | |
| | 20 | Very sick; hospital admission necessary; active supportive treatment necessary | |
| | 10 | Moribund; fatal processes progressing rapidly | |
| | 0 | Dead | |

Supplemental Figure 8 **IBTP Demographic and Follow Up Forms** | Standardized forms completed by the nurse navigator during onboarding and at 30- and 90-day follow-up visits. These forms document demographic information, quality-of-life metrics, clinical concerns, and treatment planning. They also include Karnofsky performance status scoring and track outstanding imaging or laboratory investigations. These structured assessments enable early detection of clinical deterioration and ensure timely adjustments to care plans through multidisciplinary coordination.

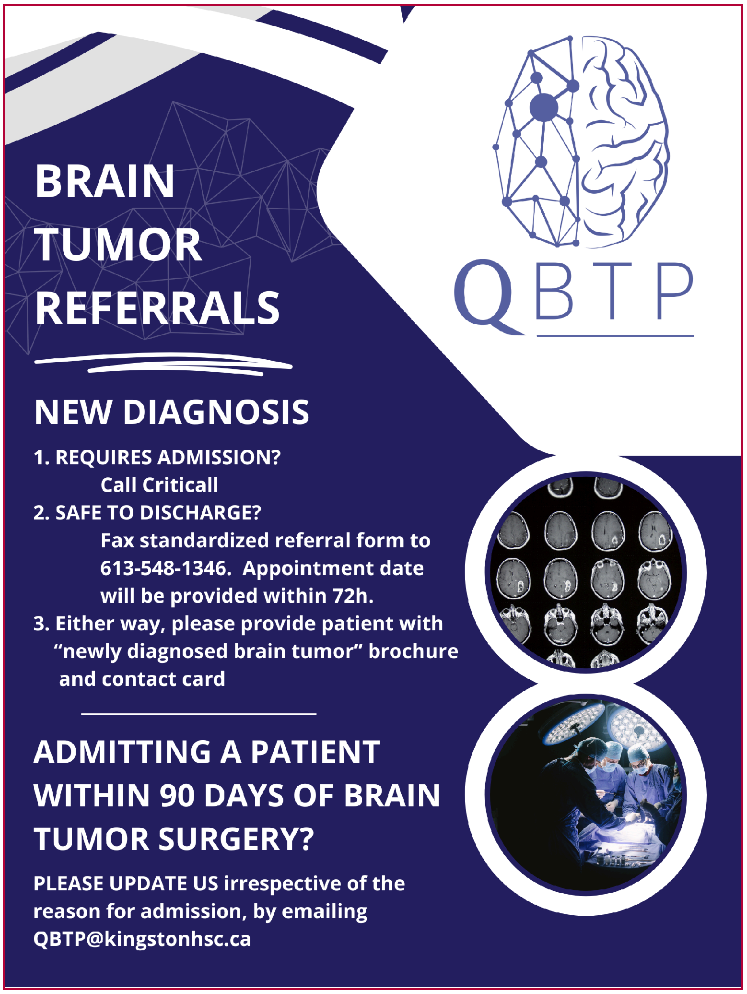

Supplemental Figure 9 **IBTP Offsite Healthcare Poster** | Poster distributed to regional hospitals and community providers to guide the referral process for patients with a new brain tumour diagnosis. It outlines steps for urgent and non-urgent admissions, use of the standardized referral form, and distribution of educational materials. The poster also instructs providers to notify the IBTP team of any readmissions within 90 days of surgery to ensure care continuity and capture outcomes data.

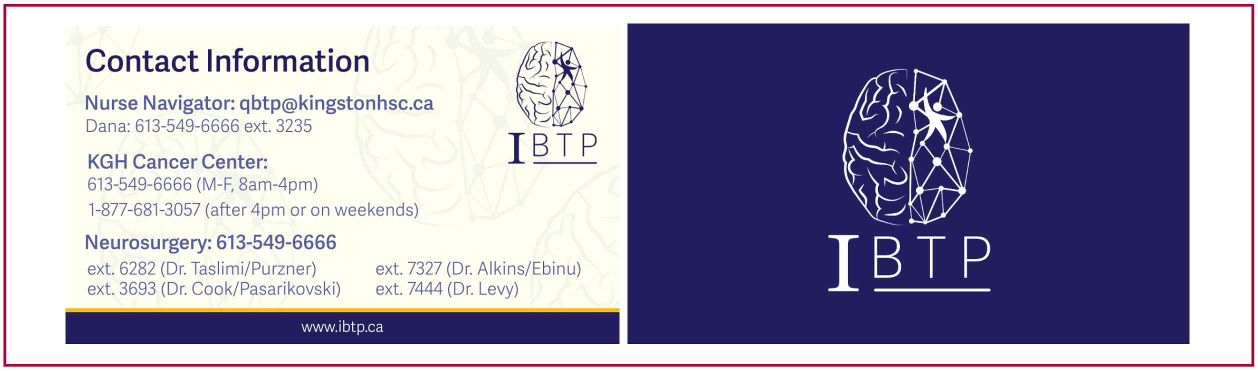

Supplemental Figure 10 **IBTP Contact Card** | Contact information cards distributed to each patient for timeliness of access. The cards include contact information for neurosurgery, the cancer clinic, and the Nurse Navigator. Patients were given instructions on the best touch point for their inquiries, questions, and concerns.

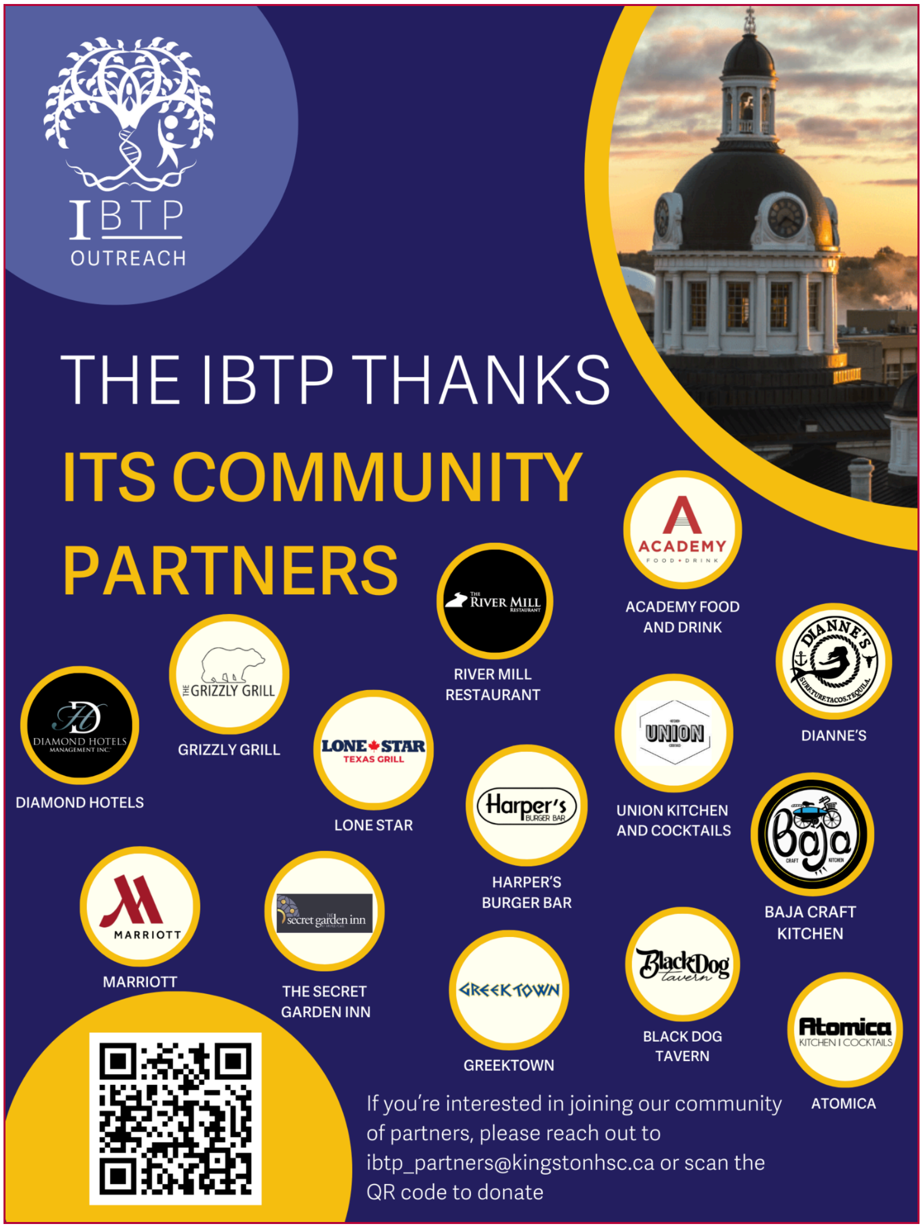

Supplemental Figure 11 **IBTP Community Partners** | Poster designed to inform patients, families, and caregivers of the community partners program available through the IBTP. Patients and their families are offered subsidized food and accommodations options for the duration of their treatment stay to reduce cost barriers to long treatment courses.

QBTP

### PATIENT SATISFACTION SURVEY

**TO BE COMPLETED BY NURSE:**

**DATE COMPLETED:** ______________________
(DD/MM/YYYY)

**RESEARCH ID:** ______________________

**WHAT IS QBTP?** The Queen's Brain Tumor Program (QBTP) was established to address the critical and urgent need for equitable, high-quality healthcare for individuals diagnosed with brain tumors in South-Eastern Ontario. QBTP is committed to supporting patients and their loved ones as they travel through each phase of their brain tumor journey.

**WHAT IS THIS SURVEY FOR?** If you would be willing to complete the following survey, your input would **provide valuable insight from a patient perspective** that allows us to better understand the needs of our patients and adapt to better meet these needs moving forward. Please feel free to provide as much or as little feedback as you feel comfortable with. *All feedback will be kept de-identified* and will **only** be utilized for the purpose of further improving clinical care for patients diagnosed with brain tumors. Thank you in advance for your help!

***PLEASE CIRCLE THE NUMBER THAT BEST DESCRIBES YOUR EXPERIENCE WITH THE QBTP:***

| | **STRONGLY DISAGREE** | | | | **NEUTRAL** | | | | | **STRONGLY AGREE** |
|---|---|---|---|---|---|---|---|---|---|---|
| The QBTP helped me effectively navigate my treatment. | 1 | 2 | 3 | 4 | 5 | 6 | 7 | 8 | 9 | 10 |
| The QBTP effectively met my individual needs as a patient. | 1 | 2 | 3 | 4 | 5 | 6 | 7 | 8 | 9 | 10 |
| I would recommend QBTP to someone I know if they were diagnosed with a brain tumor. | 1 | 2 | 3 | 4 | 5 | 6 | 7 | 8 | 9 | 10 |

***PLEASE CIRCLE THE NUMBER THAT BEST DESCRIBES HOW SATISFIED YOU ARE WITH THE FOLLOWING ASPECTS OF QBTP BASED ON YOUR EXPERIENCE:***

| | **VERY UNSATISFIED** | | | | **NEUTRAL** | | | | | **VERY SATISFIED** |
|---|---|---|---|---|---|---|---|---|---|---|
| **Patient Information Materials**<br>*(e.g., brochures, website, etc.)* | 1 | 2 | 3 | 4 | 5 | 6 | 7 | 8 | 9 | 10 |
| | Did you receive any of our patient information materials?<br>*(e.g., patient information package, a QBTP brochure, QR code or website URL.)* | | | | | | ☐ YES | | | ☐ NO |
| **Timeliness of Care**<br>*(e.g., wait times between surgery & oncology)* | 1 | 2 | 3 | 4 | 5 | 6 | 7 | 8 | 9 | 10 |
| **Patient Centric Care**<br>*(e.g., feeling supported, heard & valued by QBTP/ your care team)* | 1 | 2 | 3 | 4 | 5 | 6 | 7 | 8 | 9 | 10 |

What has been the best/most helpful part of your experience so far with the Queen's Brain Tumor Program (QBTP)?
*(e.g., educational resources, nurse navigator, website, etc.)*

______________________________________________

What is something you think QBTP could improve upon or change to better meet the needs of patients diagnosed with brain tumors?

______________________________________________

Do you have any other concerns, challenges or feedback that you would like us to know about?
*(e.g. living far away from hospital, financial concerns, future planning, anxiety, etc.)*

______________________________________________

Supplemental Figure 12 **IBTP Patient Survey|** Satisfaction survey sent to patients to obtain perceived care quality and program feedback. Sent to all patients at their 30 and 90 day visits to inform iterative program changes.

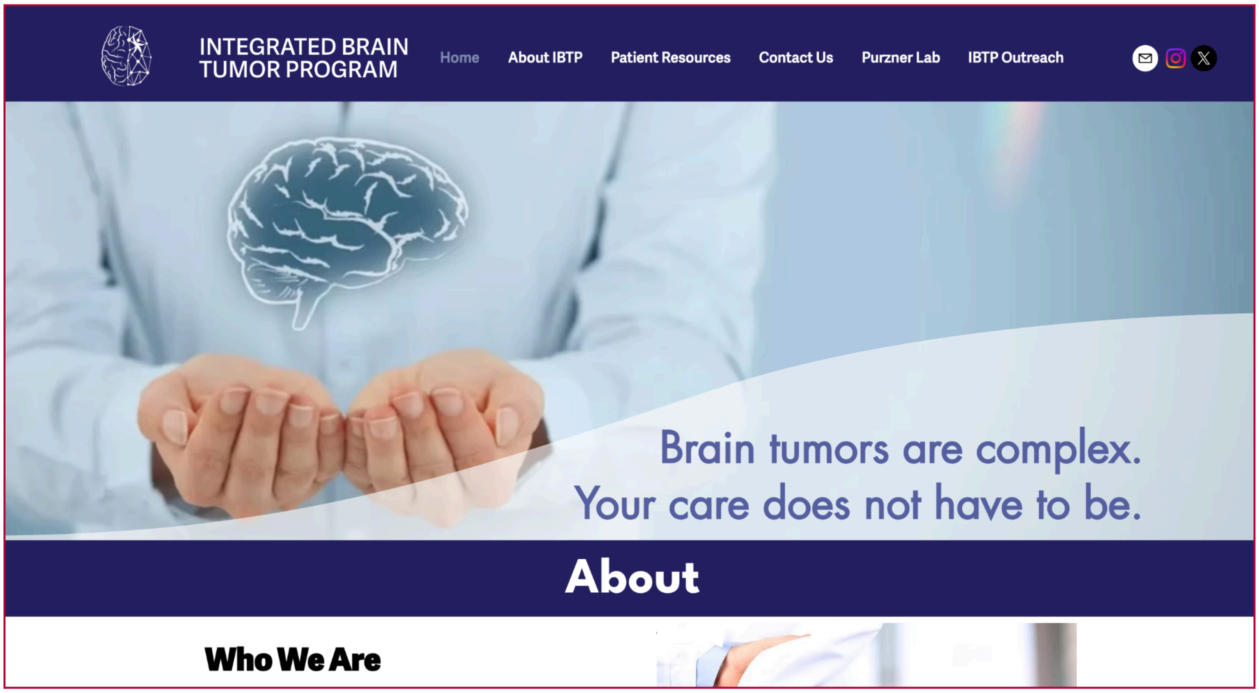

Supplemental Figure 13 **IBTP Website** | Introduction page to public IBTP website. Website tracking metrics provide insight to search interest and brand interactions.

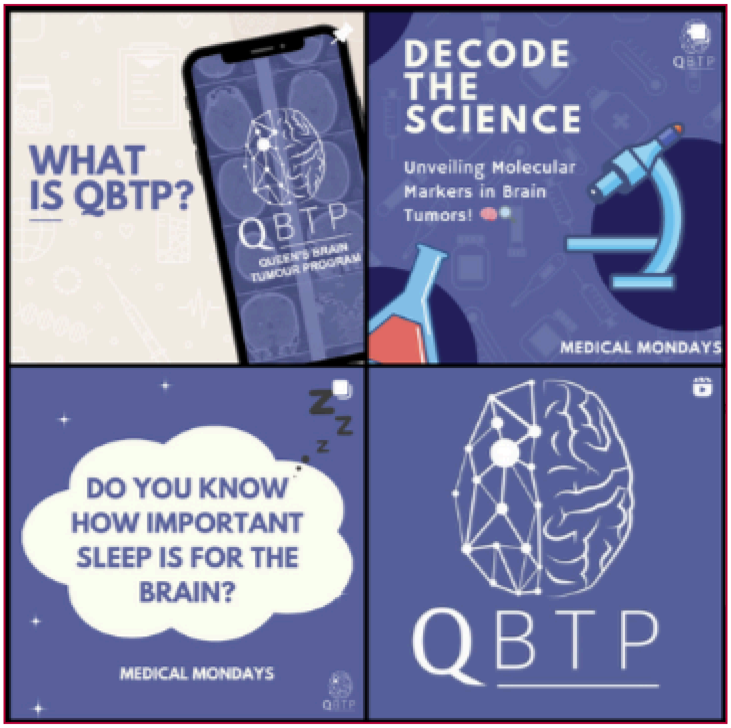

Supplemental Figure 14 **IBTP Social Media Instagram** | IBTP Instagram social media account with regular educational posting for increased program awareness and media presence.

#### Supplemental Figures

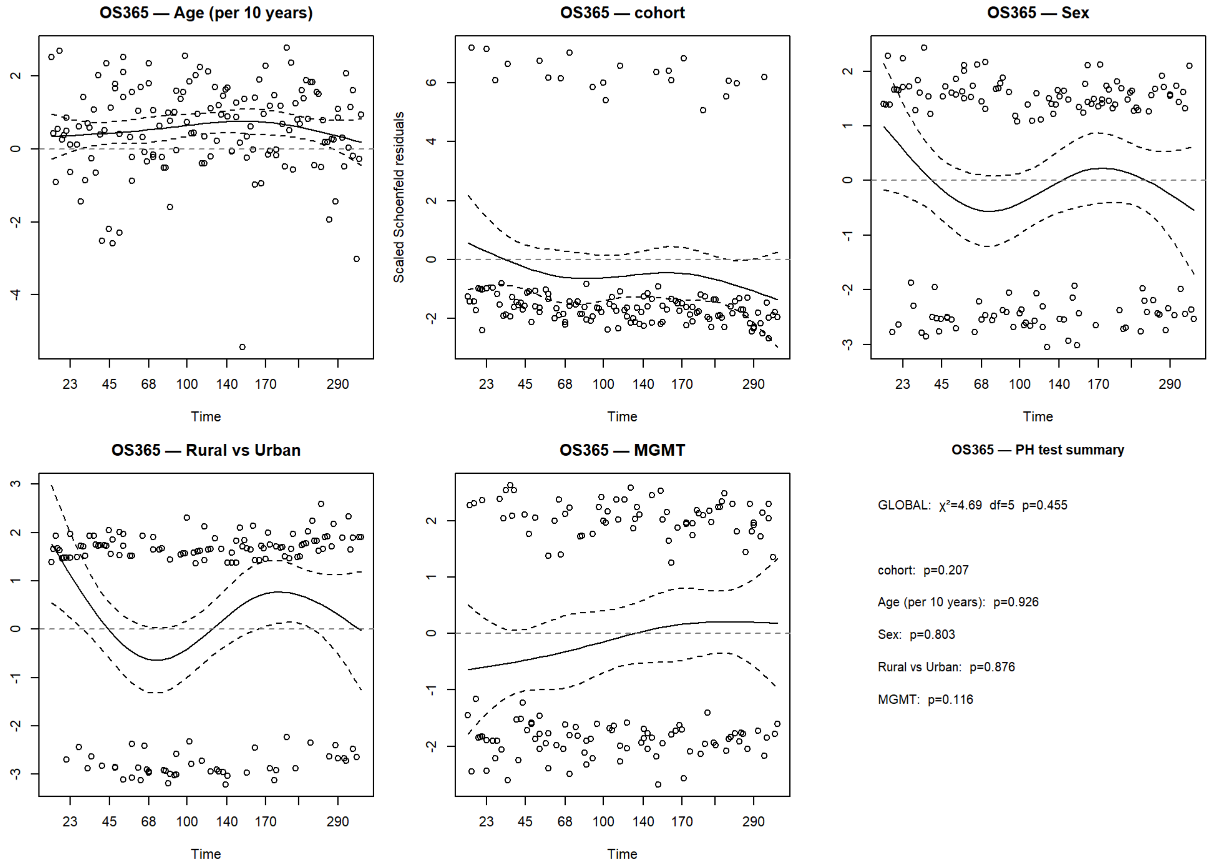

Supplemental Figure 15 **Schoenfeld Residual Plots for One Year Survival** | Scaled Schoenfeld residual plots assessing proportional hazards assumptions for covariates included in the Cox proportional hazards model for one-year overall survival. No statistically significant deviations from proportionality were detected for cohort, age, sex, or rurality.

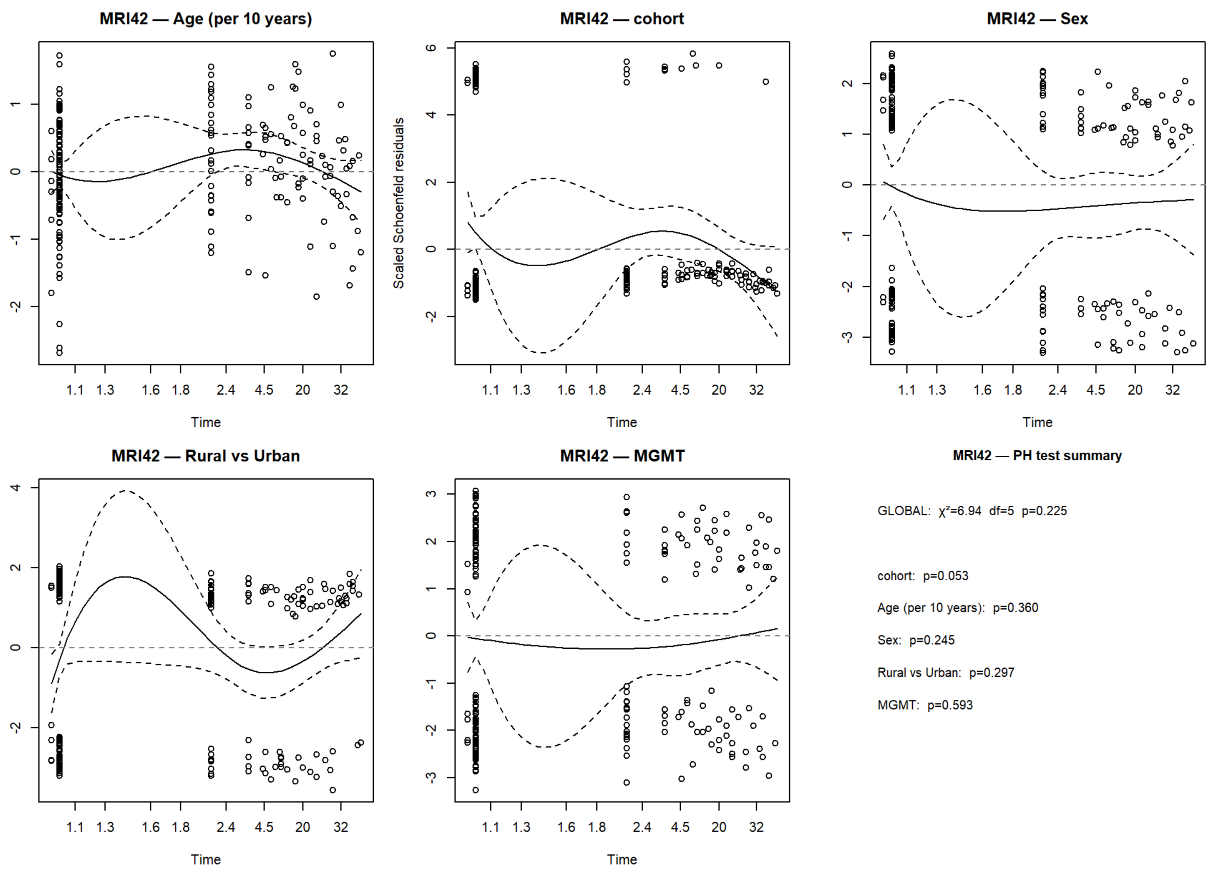

Supplemental Figure 16 **Schoenfeld Residual Plots for Postoperative MRI** | Scaled Schoenfeld residual plots assessing proportional hazards assumptions for covariates included in Cox proportional hazards models evaluating postoperative MRI within 48 hours. A time-dependent effect was observed for cohort status, motivating complementary competing-risk and sensitivity analyses; other covariates satisfied proportional hazards assumptions.

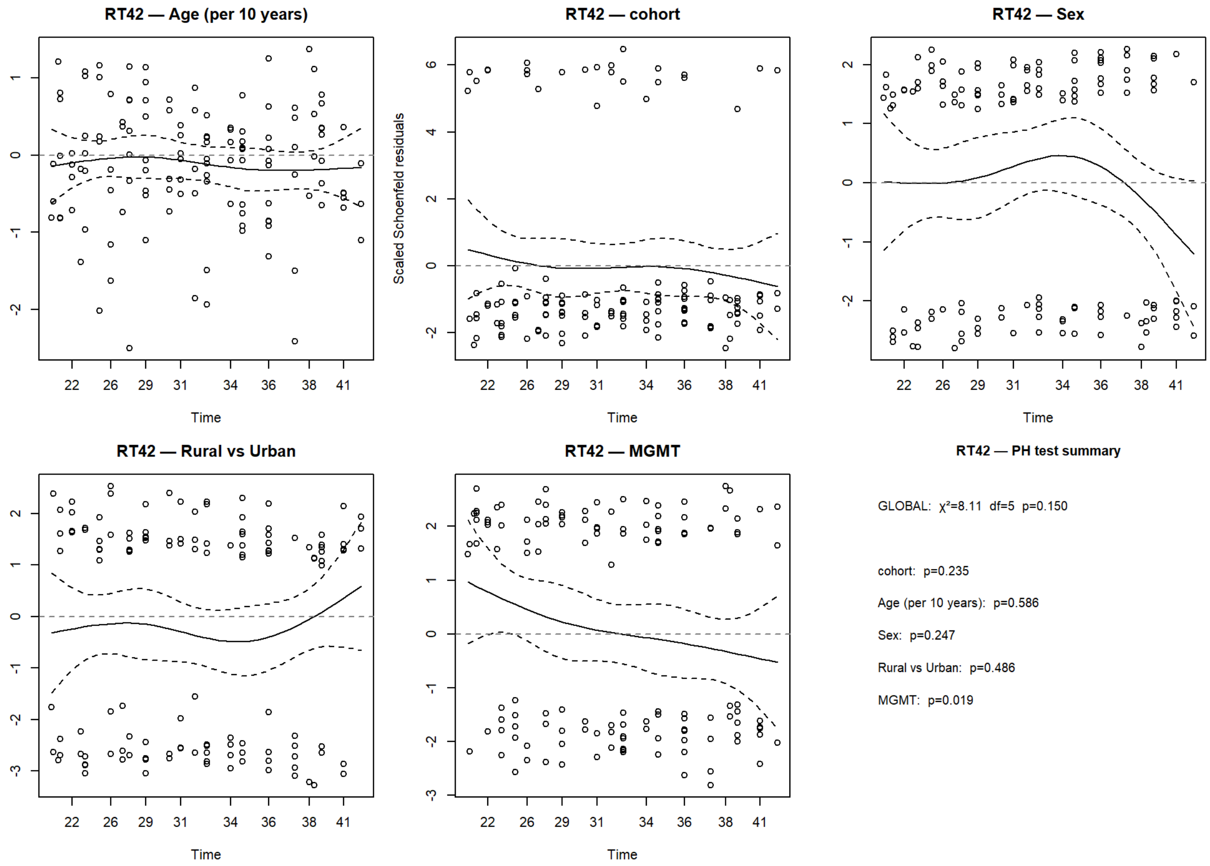

Supplemental Figure 17 **Schoenfeld Residual Plots for Radiotherapy Start** | Scaled Schoenfeld residual plots assessing proportional hazards assumptions for covariates included in Cox proportional hazards models evaluating initiation of radiotherapy within six weeks of surgery. No violations of the proportional hazards assumption were detected.

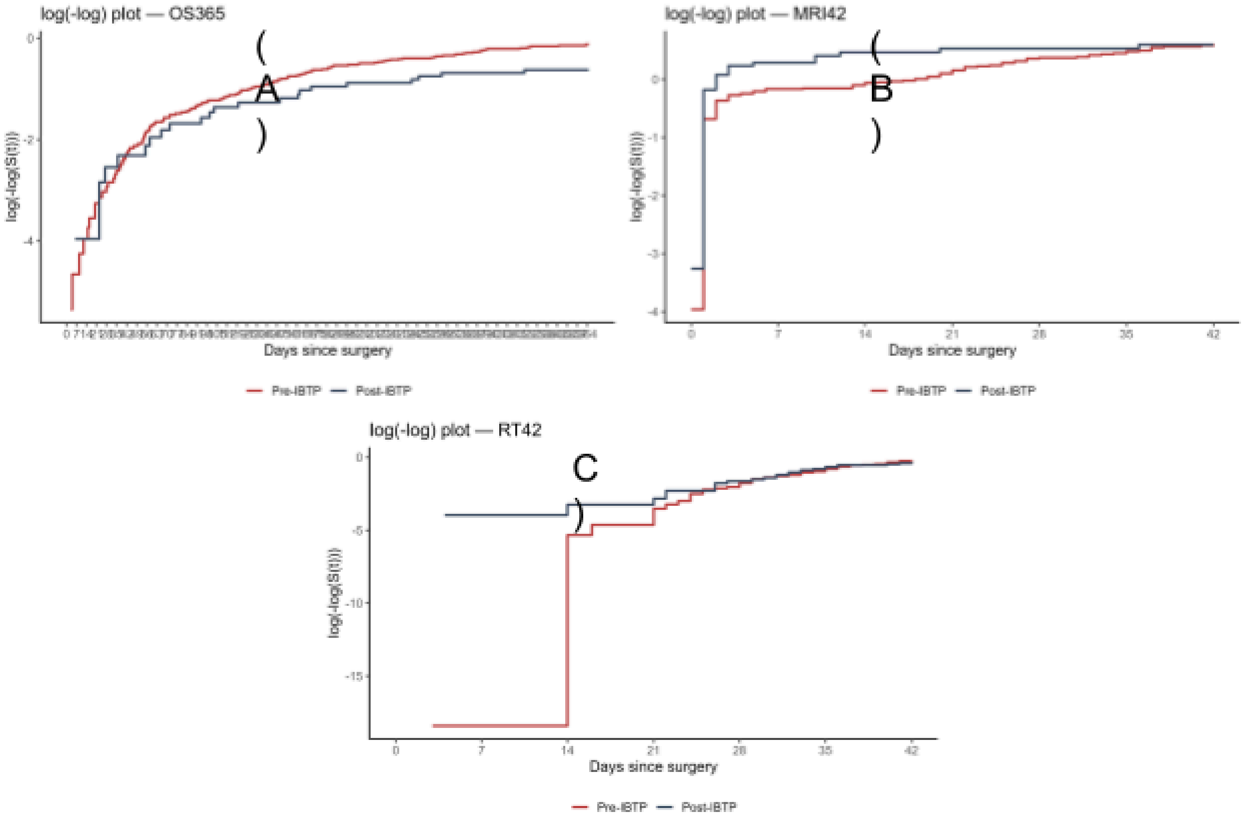

Supplemental Figure 18 **Log(-log) Survival Plots** | Log(−log) survival plots stratified by cohort for (A) one year overall survival; (B) postoperative MRI timeliness; and (C) radiotherapy initiation. Approximate parallelism of curves supports the proportional hazards assumption across endpoints following final model specification.

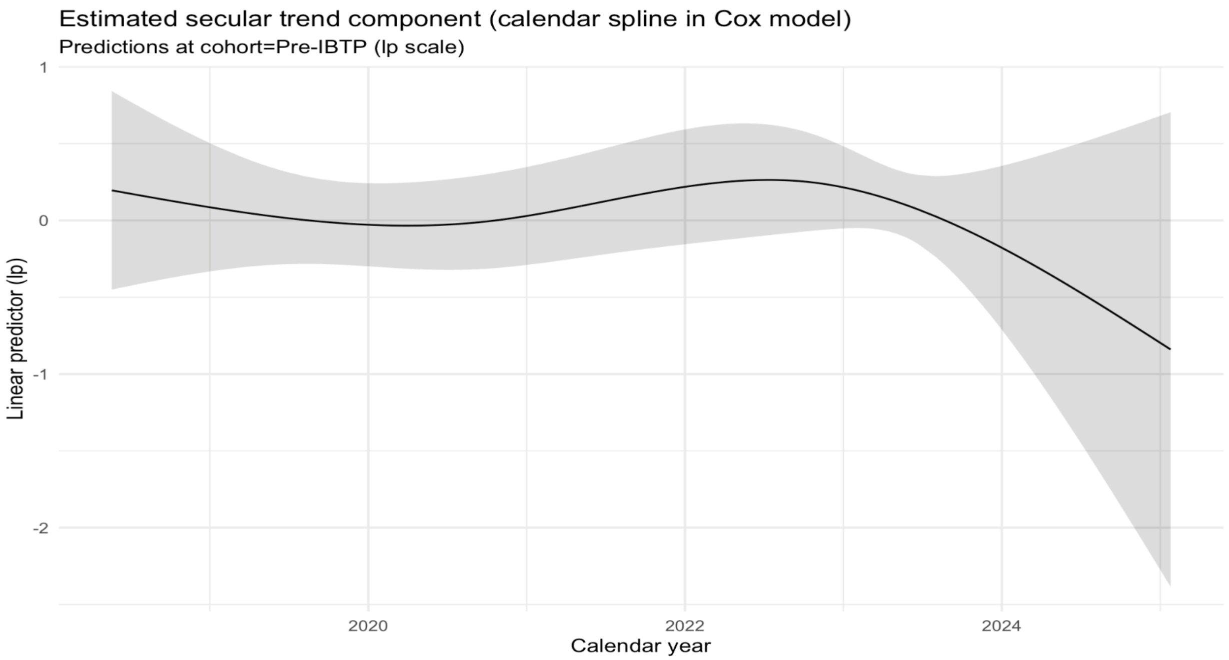

| df <int> | AIC <dbl> | HR <dbl> | LCL <dbl> | UCL <dbl> | p <dbl> |
|---|---|---|---|---|---|
| 1 | 1732.926 | 0.5493107 | 0.3033907 | 0.9945667 | 0.04793417 |
| 2 | 1734.882 | 0.5886620 | 0.2359576 | 1.4685814 | 0.25592920 |
| 3 | 1734.794 | 1.0610021 | 0.2951766 | 3.8137356 | 0.92772114 |
| 4 | 1734.919 | 1.4290864 | 0.4234683 | 4.8227642 | 0.56507053 |
| 5 | 1734.535 | 1.3607928 | 0.4487684 | 4.1263091 | 0.58623470 |

5 rows

Supplemental Figure 19 **Calendar-Time Secular Trend Modeling**| Estimated secular trend component from Cox models incorporating restricted cubic splines for calendar year. The upper panel displays the modeled linear predictor with 95% confidence bands. The lower panel summarizes model fit across spline degrees of freedom, demonstrating minimal improvement beyond low-order specifications.

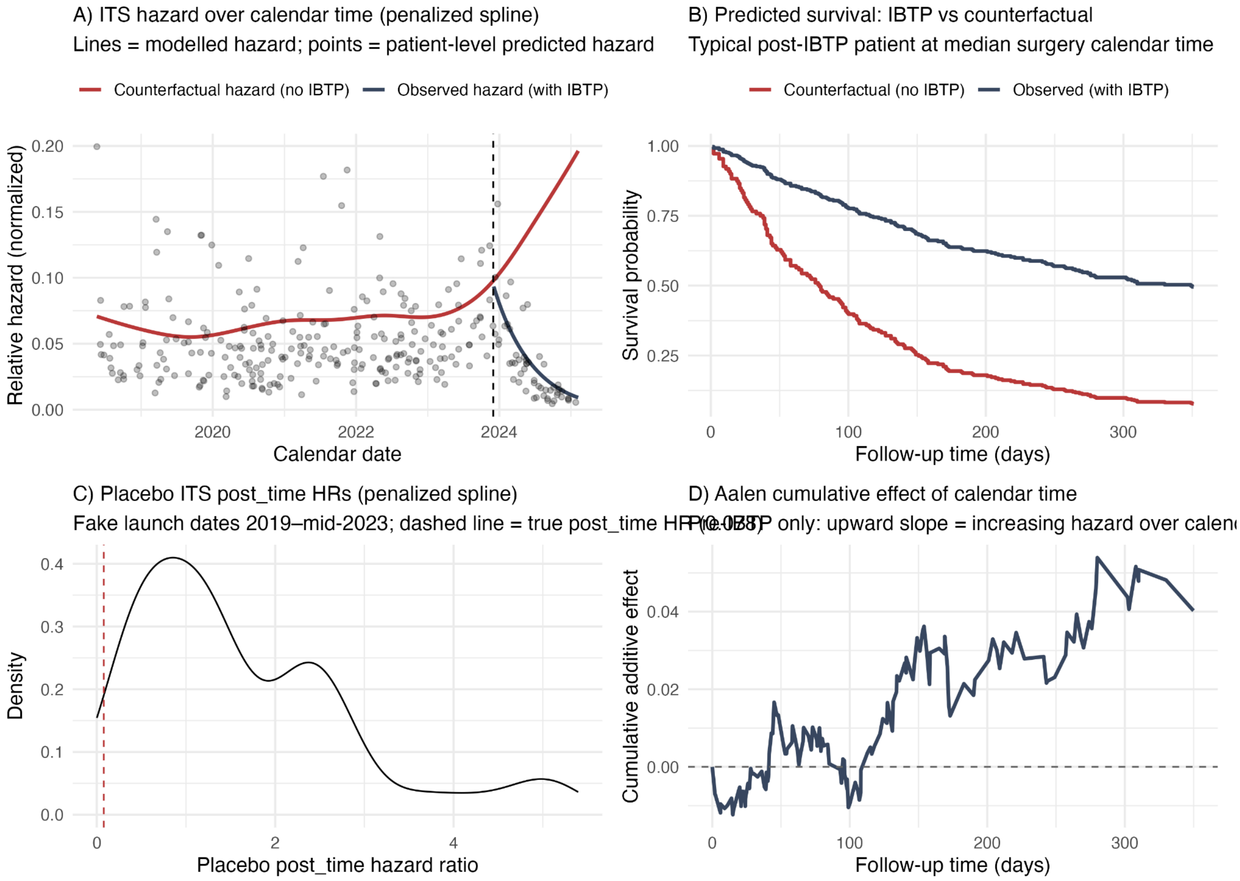

Supplemental Figure 20 **Placebo Interrupted Time Series Analyses Test** | Results of placebo interrupted time series analyses evaluating the specificity of the observed post-implementation effect. Panels depict (A) modeled hazard over calendar time, (B) predicted survival under observed versus counterfactual scenarios, (C) distribution of placebo post-implementation hazard ratios using false intervention dates, and (D) cumulative hazard tests, supporting temporal specificity of the observed effect.

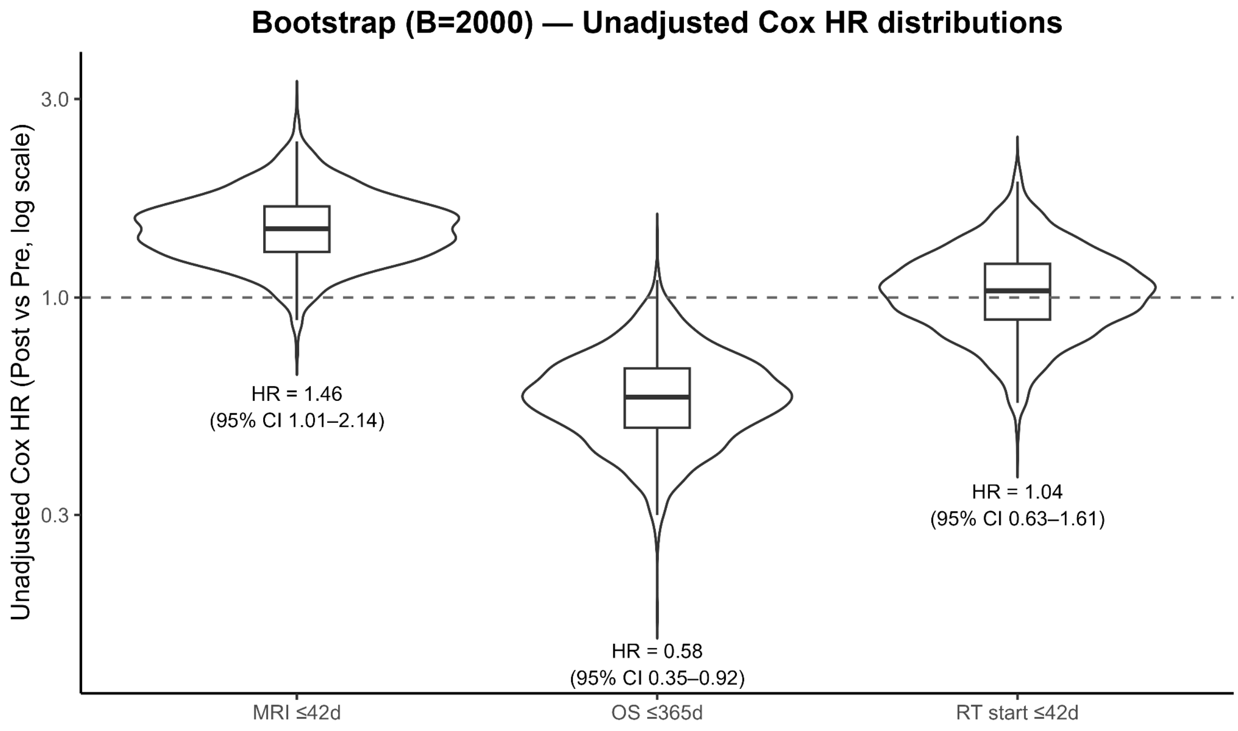

Supplemental Figure 21 **Bootstrap Analyses Across Endpoints**| Bootstrap distributions of hazard ratios for postoperative MRI timeliness, radiotherapy initiation, and one-year overall survival. Density plots and percentile-based confidence intervals demonstrate stability of effect estimates across resampled datasets.

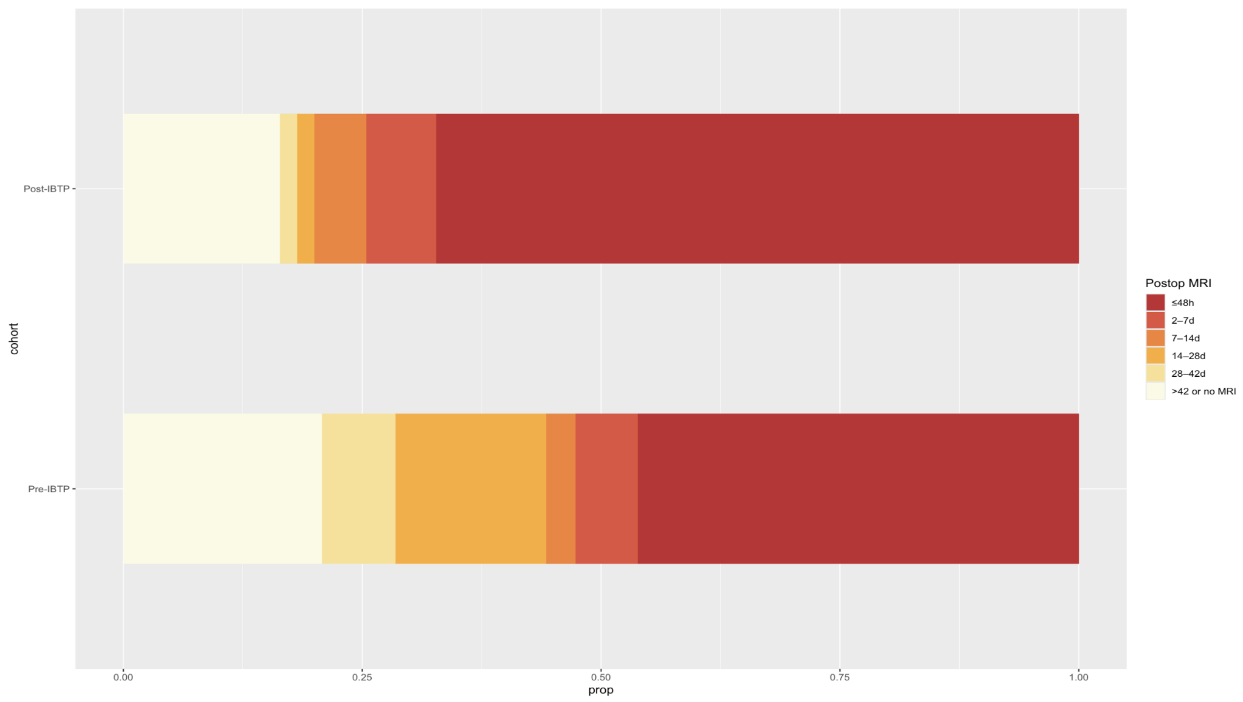

Supplemental Figure 22 **Distribution of Postoperative MRI Timing By Cohort**| Thermal plot illustrating the distribution of postoperative MRI timing intervals by cohort. Color intensity reflects the proportion of patients within predefined postoperative time bins, highlighting shifts toward earlier imaging following program implementation.
